# The UK Biobank submaximal cycle ergometer test for assessment of cardiorespiratory fitness: Validity, reliability, and association with disease outcomes

**DOI:** 10.1101/2020.09.29.20203828

**Authors:** Tomas I. Gonzales, Kate Westgate, Tessa Strain, Stefanie Hollidge, Justin Jeon, Dirk L. Christensen, Jorgen Jensen, Nicholas J. Wareham, Søren Brage

**Affiliations:** MRC Epidemiology Unit, University of Cambridge, Cambridge, United Kingdom; University of Copenhagen, Denmark; University of Oslo, Norway

## Abstract

**Background:** Cardiorespiratory fitness (CRF) was assessed in UK Biobank (UKB) using heart rate response to a submaximal ramped cycle ergometer test that was individualised for participant characteristics including cardiovascular disease risk. Studies have since explored health associations with CRF by estimating maximal oxygen consumption (VO_2_max) from UKB test data using interpretation methods that have not accounted for this individualisation procedure. Thus, dose-response relationships reported in these studies may be inaccurate. We developed and validated a novel VO_2_max estimation approach that accounts for the UKB test individualisation procedure and compared dose-response relationships with health outcomes between the novel and previous methods.

**Methods:** In a cross-over study (n=189), participants completed several UKB tests and VO_2_max was measured. A multilevel modelling framework was developed that combines heart rate response features from the UKB test to estimate VO_2_max. Estimates were compared within participants across UKB test protocols, and with directly measured VO_2_max. Short-term test-retest reliability was assessed in a subsample of participants (n=87). In UKB, we examined associations between estimated CRF and disease endpoints (n=80,259) and compared associations obtained with an unvalidated method. Long-term test-retest reliability was examined (n = 2877).

**Results:** Estimated and directly measured VO_2_max were strongly correlated (Pearson’s *r* range: 0.68 to 0.74) with no mean bias (women bias: −0.8 to 0.4; men bias range: −0.3 to 0.3), outperforming a previous approach for interpreting UKB test data. Agreement between estimated VO_2_max across different test protocols was strong (Pearson’s *r* range: 0.94 to 0.99). Short- and long-term reliability was also high (lambda=0.91 and 0.80, respectively). All-cause mortality was 7% (95%CI 4-10%, 2686 deaths) lower and CVD mortality 9% (95%CI 3-14%, 858 deaths) lower for every 1-MET difference in fitness, associations twice as strong as determined by previous methods.

**Conclusions:** We present a valid and reliable method for estimating CRF in UKB and demonstrate its utility in characterising dose-response relationships with health outcomes. Accounting for the individualisation procedure strengthens observed relationships between CRF and disease and enhances the case for promoting improved fitness in the general population.

## Introduction

Maximal oxygen consumption (VO_2_max) is a powerful predictor of all-cause and cause-specific mortality ^1–3^ and morbidity ^4–7^ but is rarely directly measured in large-scale population-based studies due to cost and safety concerns ^8^. As an alternative, a variety of methods have been developed to indirectly measure VO_2_max from heart rate (HR) response to incremental submaximal cardiorespiratory fitness (CRF) tests ^9^. To ensure participant safety, screening procedures are generally put in place to exclude individuals from CRF testing which potentially leads to selection bias. Somewhat ironically, it is the excluded participants that are more likely to experience incident disease events following a baseline assessment, thus making it more difficult to examine the relationship between CRF and these diseases in epidemiological studies with such designs.

The exercise tests typically employed in population studies are broadly classified as either steady-state tests or ramped tests. Steady-state tests consist of several stepwise work rate (WR) increments every 4-6 minutes, allowing time for HR and VO_2_ to stabilise at each WR. Methods for estimating VO_2_max from submaximal HR responses to incremental steady-state WR have been well-studied and validated ^10^. Steady-state testing, however, can be long and inefficient depending on the number of WR increments and is impractical for populations with low exercise tolerance. The alternative is to use ramped tests, where WR is increased constantly and continuously in small increments. This allows HR and VO_2_ response to be characterised over a wider range of WR values in less time and enables the rate at which WR is increased (i.e. ramp rate) to be individualised to the participant’s ability and contra-indications to exercise. With these practical advantages, however, come several methodological issues. At a given ramped WR, HR and VO_2_ values will be less than those measured during a steady-state test at an equivalent WR ^11,12^. Thus, VO_2_max estimates from ramped tests may be biased if the HR- or VO_2_-ramp response is extrapolated from submaximal to maximal levels using methods validated for steady-state tests. Several studies provide alternative methods for estimating VO_2_max from HR- or VO_2_-ramp response ^13–15^. These methods may be valid for ramped tests at common ramp rates but are insufficient for tests individualised across a wide range of ramp rates.

The UK Biobank (UKB), a prospective cohort study of over half a million UK residents, used a ramped submaximal cycle ergometer test (henceforth referred to as the “UKB CRF test”) to measure CRF in a subsample of 100,000 participants. The UKB CRF test was designed to be as inclusive as possible; tests were short, had relatively low work rates, and were individualised depending on both presumed ability (from body size and resting HR measures) and a preliminary health risk assessment, resulting in 22 protocols (11 for men and women each) with different initial work rates and ramp rates. This strategy allowed testing to be conducted safely across participants with a wide fitness range while including those not normally considered for exercise testing, minimising the issue of test data only being available in those less likely to experience disease outcomes. No adverse events attributed to acute exercise testing were observed. Previous attempts at estimating VO_2_max from UKB CRF test data have relied on methods assuming no difference in VO_2_max estimation bias between tests with different ramp rates or that only utilise a small proportion of available test data ^16–22^. While these approaches may broadly rank individuals by fitness level, their validity against gold-standard CRF measures is unknown which leads to challenges in interpreting epidemiological findings, in particular characterising dose-response relationships between CRF and disease endpoints. Differential VO_2_max estimation bias across tests would lead to attenuation of observed relationships, an issue which is exacerbated for methods that do not use the totality of the HR response since measurement noise would disproportionately influence CRF estimates.

In this study, we develop and validate a novel VO_2_max estimation method for the UKB CRF test using exercise test data from a validation study of participants, age-, sex- and BMI-matched to the UKB sample. We first introduce the modelling framework for our method: the features to be extracted from the HR response to exercise and how those features will be combined in a multilevel estimation model. We use our method to estimate maximal WR and VO_2_max values, and evaluate estimation model performance against directly measured VO_2_max from an independent test in the validation study participants. We then apply our method to exercise test data from UKB cohort participants and use survival analyses to examine CRF-disease associations. Finally, we compare our findings with previous investigations of CRF in the UKB.

## Methods

### Validation of UKB CRF test

#### Validation study participants

We recruited a subsample of participants from the Fenland study, a population-based study in Cambridgeshire, UK ^23^, using a stratified random sampling procedure (Supplemental Table 1). Exclusion criteria were: heart pacemaker; unable to walk without aid; history of angina pectoris; blood pressure greater than 180/110 mm Hg; musculoskeletal injury that would impair cycling on the ergometer; pregnancy; and currently taking cardioactive drugs (e.g. beta-blockers, aspirin). Ethical approval was obtained by the University of Cambridge Human Biology Research Ethics Committee (Ref: HBREC/2015.16). All participants provided written informed consent.

**Table 1.** Validation study participant characteristics.

| Characteristics | Women (n = 86) | Men (n = 105) |
| --- | --- | --- |
| Age (y) | 55.0 ± 6.5 | 54.3 ± 7.3 |
| Height (cm) | 164.0 ± 6.4 | 178.5 ± 6.7 |
| Weight (kg) | 69.2 ± 10.8 | 84.8 ± 10.2 |
| BMI (kg · m<sup>-2</sup>) | 25.7 ± 3.4 | 26.6 ± 3.0 |
| RHR (bpm) | 61.3 ± 8.4 | 61.4 ± 10.4 |
| WR at LT (W) * | 103.4 ± 15.0 | 162.2 ± 23.5 |
| WR at RCP (W) * | 140.8 ± 25.4 | 227.0 ± 40.0 |
| WRmax (W) * | 158.5 ± 28.9 | 257.0 ± 55.5 |
| HRmax (bpm) * | 168.4 ± 10.7 | 169.8 ± 14.0 |
| VO<sub>2</sub>max (ml · min<sup>-1</sup> · kg<sup>-1</sup>) † | 30.2 ± 5.8 | 36.3 ± 6.1 |

BMI: Body mass index, RHR: Resting heart rate, WR: Work rate, LT: Lactate threshold, RCP: Respiratory compensation point, WRmax: Measured maximal work rate, HRmax: Measured maximal heart rate, VO<sub>2</sub>max: Maximal Oxygen consumption. \*: Characteristics computed in participant subsample with valid maximal test data (Women: n = 77; Men: n = 89). †: Characteristics computed in participants who reached VO<sub>2</sub>max (Women: n = 55; Men: n = 78).

#### Experimental procedure and equipment

Validation study participants were screened according to standardised procedures used for the UKB CRF test ^24^ (see Supplemental Materials for a description of the UKB CRF protocols). Then, participants completed the UKB flat test, two UKB ramped tests at different ramp rates, a steady-state test (unique to the validation study), and another ramped test (validation only) to elicit VO_2_max (Figure 1A). Tests were conducted consecutively, separated by at least 15 minutes of rest, and were specified according to the test that the participant would have been assigned had s/he been part of UKB (see Supplemental Table 2). The target (highest) WR for the second ramped test was at least 30W greater than the first; thus, each participant completed a “low” and “high” ramped UKB test. The steady-state test consisted of four incremental 4-minute flat-phases with each WR increment ranging from 10-20W. For the ramped max test, participants were fitted with a face mask to measure respiratory ventilation and gas exchange and cycled while WR increased until exhaustion. VO_2_max was reached if two of the following criteria were met: a respiratory exchange ratio exceeding 1.20; no VO_2_ increase despite increasing WR (< 2.5 ml O_2_ · kg^-1^ · min^-1^); and no HR increase despite increasing WR. VO_2_max was measured as the average of the two highest VO_2_ measurements in the last forty-five seconds of the test. WR values were measured at exhaustion (i.e. maximal work rate achieved on the test), the lactate threshold (LT), and at the respiratory compensation point (RCP; see Supplemental Methods).

**Table 2.** UKB participant characteristics across CRF tertiles.

| Sex | Women |  |  |  |  |  | Men |  |  |  |  |  |
| --- | --- | --- | --- | --- | --- | --- | --- | --- | --- | --- | --- | --- |
|  | Lower |  | Middle |  | Higher |  | Lower |  | Middle |  | Higher |  |
|  | N | VO<sub>2</sub>max | N | VO<sub>2</sub>max | N | VO<sub>2</sub>max | N | VO<sub>2</sub>max | N | VO<sub>2</sub>max | N | VO<sub>2</sub>max |
| CRF Tertiles |  |  |  |  |  |  |  |  |  |  |  |  |
| Age stratum (y) |  |  |  |  |  |  |  |  |  |  |  |  |
| Younger than 50 | 3037 | 21.4 ± 2.7 | 3037 | 27.2 ± 1.3 | 3037 | 33.6 ± 3.4 | 2542 | 28.0 ± 2.7 | 2543 | 34.0 ± 1.5 | 2542 | 40.8 ± 3.4 |
| 50-54 | 2178 | 20.8 ± 2.5 | 2180 | 26.2 ± 1.3 | 2178 | 32.5 ± 3.4 | 1687 | 27.1 ± 2.7 | 1688 | 33.0 ± 1.5 | 1687 | 39.9 ± 3.5 |
| 55-59 | 2542 | 20.3 ± 2.4 | 2543 | 25.4 ± 1.2 | 2542 | 31.4 ± 3.3 | 2004 | 26.6 ± 2.6 | 2005 | 32.4 ± 1.4 | 2004 | 38.9 ± 3.5 |
| 60-64 | 3497 | 19.6 ± 2.3 | 3498 | 24.3 ± 1.1 | 3497 | 30.0 ± 3.3 | 3096 | 26.0 ± 2.5 | 3096 | 31.7 ± 1.3 | 3096 | 37.9 ± 3.3 |
| 65 and older | 2892 | 19.0 ± 2.3 | 2893 | 23.5 ± 1.1 | 2892 | 28.8 ± 3.1 | 3244 | 25.3 ± 2.4 | 3245 | 30.7 ± 1.3 | 3244 | 36.9 ± 3.4 |
| Combined across age stratum | 14146 | 20.2 ± 2.6 | 14151 | 25.2 ± 1.8 | 14146 | 31.2 ± 3.7 | 12573 | 26.5 ± 2.7 | 12577 | 32.2 ± 1.8 | 12573 | 38.6 ± 3.7 |
| Age (y) |  | 57.1 ± 8.0 |  | 57.1 ± 8.0 |  | 56.9 ± 8.1 |  | 57.9 ± 8.3 |  | 57.9 ± 8.3 |  | 57.8 ± 8.4 |
| Height (cm) |  | 162.5 ± 6.3 |  | 162.9 ± 6.2 |  | 162.9 ± 6.2 |  | 176.3 ± 6.8 |  | 175.9 ± 6.7 |  | 175.3 ± 6.7 |
| Weight (kg) |  | 79.2 ± 14.7 |  | 69.6 ± 10.2 |  | 62.8 ± 8.5 |  | 94.0 ± 14.3 |  | 84.7 ± 10.9 |  | 77.4 ± 9.8 |
| Body mass index |  | 30.0 ± 5.3 |  | 26.2 ± 3.5 |  | 23.7 ± 3.0 |  | 30.2 ± 4.2 |  | 27.3 ± 3.0 |  | 25.2 ± 2.8 |
| Fat-free mass (kg) |  | 46.2 ± 5.4 |  | 43.9 ± 4.3 |  | 42.5 ± 3.9 |  | 66.6 ± 7.9 |  | 63.1 ± 6.9 |  | 60.3 ± 6.4 |
| Resting blood pressure (mmHg) |  |  |  |  |  |  |  |  |  |  |  |  |
| Systolic |  | 134.8 ± 17.0 |  | 129.6 ± 17.1 |  | 125.6 ± 16.9 |  | 139.5 ± 15.5 |  | 135.3 ± 15.1 |  | 131.4 ± 15.0 |
| Diastolic |  | 81.7 ± 9.4 |  | 77.4 ± 8.9 |  | 74.3 ± 8.9 |  | 84.4 ± 9.3 |  | 81.0 ± 8.9 |  | 77.3 ± 8.7 |
| Resting heart rate (bpm) |  | 73.1 ± 10.1 |  | 66.5 ± 8.0 |  | 61.5 ± 7.8 |  | 73.0 ± 11.1 |  | 64.4 ± 8.5 |  | 58.0 ± 8.1 |
| FVC (L) |  | 3.0 ± 0.6 |  | 3.2 ± 0.6 |  | 3.2 ± 0.6 |  | 4.3 ± 1.0 |  | 4.4 ± 0.9 |  | 4.5 ± 0.9 |
| FEV1 (L) |  | 2.3 ± 0.5 |  | 2.4 ± 0.5 |  | 2.5 ± 0.5 |  | 3.2 ± 0.7 |  | 3.3 ± 0.7 |  | 3.4 ± 0.7 |
| PEF (L/min) |  | 335.3 ± 83.8 |  | 342.7 ± 81.1 |  | 343.1 ± 82.1 |  | 480.9 ± 122.2 |  | 491.7 ± 121.8 |  | 494.8 ± 118.1 |
| Smoking status, % |  |  |  |  |  |  |  |  |  |  |  |  |
| Never |  | 64.2% |  | 61.0% |  | 59.0% |  | 48.1% |  | 51.1% |  | 54.8% |
| Previously |  | 29.1% |  | 31.9% |  | 33.2% |  | 41.3% |  | 38.4% |  | 35.2% |
| Currently |  | 6.7% |  | 7.1% |  | 7.8% |  | 10.6% |  | 10.5% |  | 9.9% |
| Health self-rating, % |  |  |  |  |  |  |  |  |  |  |  |  |
| Excellent |  | 9.2% |  | 16.6% |  | 23.8% |  | 8.6% |  | 13.8% |  | 22.7% |
| Good |  | 61.0% |  | 65.3% |  | 62.7% |  | 54.2% |  | 62.7% |  | 60.4% |
| Fair |  | 25.7% |  | 16.0% |  | 12.0% |  | 32.1% |  | 20.7% |  | 15.0% |
| Poor |  | 3.8% |  | 1.9% |  | 1.4% |  | 4.8% |  | 2.6% |  | 1.8% |

Values are means ± standard deviations, unless otherwise indicated. CRF: Cardiorespiratory fitness, VO<sub>2</sub>max: Maximal oxygen consumption (ml · kg<sup>-1</sup> · min<sup>-1</sup>), FVC: Forced vital capacity, FEV1: Forced expiratory volume (1s), PEF: Peak expiratory flow

**Figure 1.**
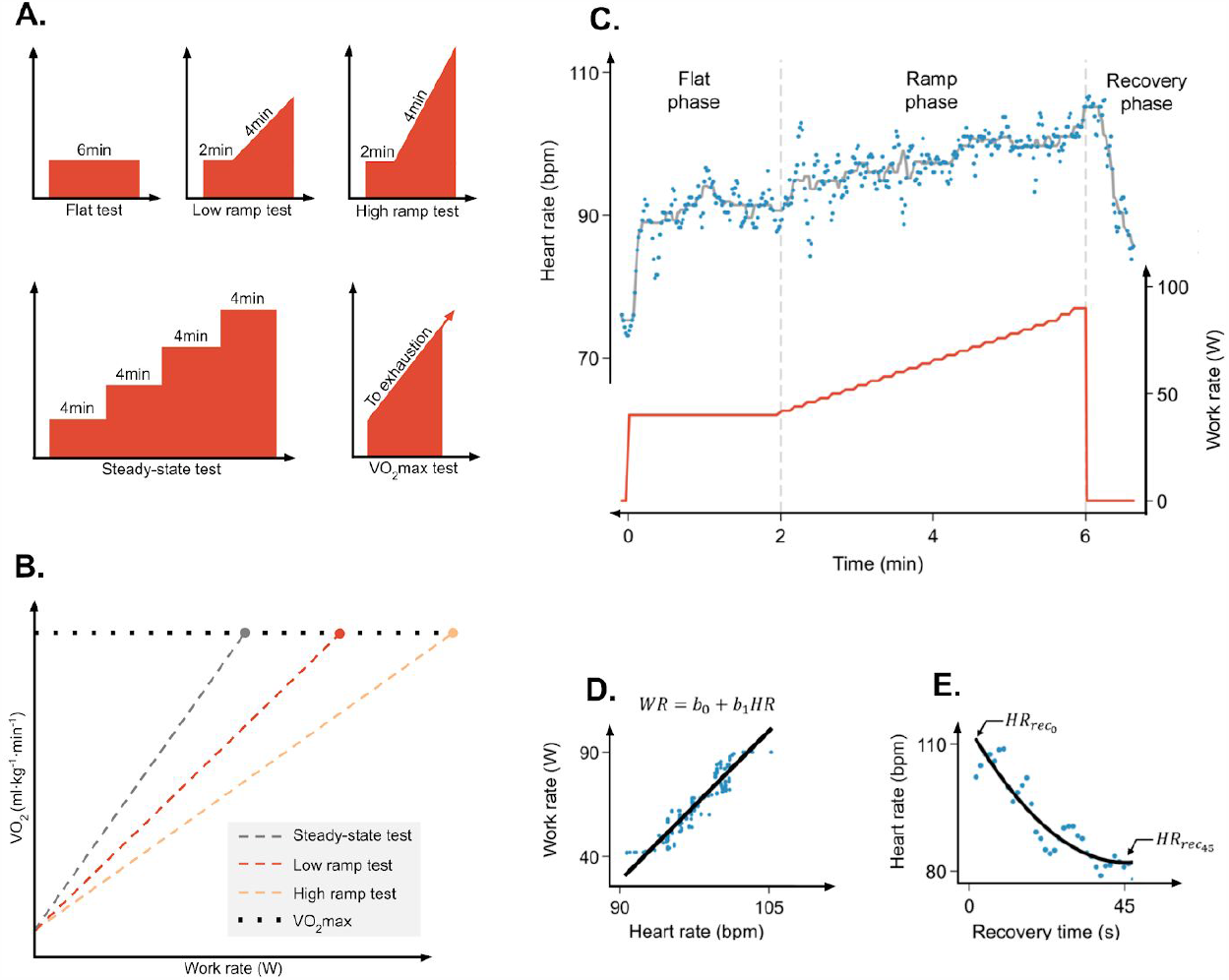
Conceptual framework and design for validation study. A. Overview of the five exercise tests performed by validation study participants (3 UKB tests, 1 steady-state and 1 max test). X-axes: Time; Y-axes: Work rate (WR). B. Conceptual plot of WR-to-VO_2_ response during steady-state and ramped exercise tests. VO_2_ increases linearly at a rate proportional to the rate of change in WR (i.e. ramp rate) until VO_2_max is reached (in an exhaustive test). The WR-to-VO_2_ relationship (line slope) changes depending on the ramp rate of the test. As ramp rate decreases, the WR when VO_2_max is achieved approaches the maximal WR for an exhaustive steady-state test. Note that VO_2_ is extrapolated to maximal values for demonstrative purposes, but in the study ramped and steady-states tests were non-exhaustive. C. Exemplar heart rate (HR) data (blue scatter & grey line; upper panel), WR data (red line; lower panel), and test phase annotation for ramp test. D. Feature extraction for ramp phase using simple linear regression model. E. Feature extraction for recovery phase using first-order exponential decay model.

Cycling was performed on an electromagnetically-braked stationary bike (eBike ergometer, GE) while electrocardiography (ECG) was recorded using 4-lead ECG (Cardiosoft) on the forearms and a Actiwave Cardio device (CamNtech, Papworth, UK) on the chest with sampling frequency of 128Hz. The 4-lead ECG leads were placed on the cubital fossa and ventral wrist of the left and right arms (mimicking the UKB protocol). Cycling work rates were controlled by computer software. Respiratory gas measurements were conducted using a computerised metabolic system with Hans Rudolph face masks (Oxycon Pro, Erich Jaeger GmbH, Hoechberg, Germany) as validated elsewhere ^25^.

All ECG signals were processed using the Physionet Toolkit implementation of the SQRS algorithm ^26^, which applies a digital filter to the signal and identifies the downward slopes of the QRS complexes ^27^. The resulting inter-beat-intervals were converted to beats-per-minute values using “ihr” of the PhysioNet Toolkit, as described previously ^16^. Pulmonary gas exchange data were sampled breath-by-breath. All data were linearly interpolated to derive quasi-continuous HR response and respiratory measures at 1s time resolution.

#### Conceptual and modeling framework for VO_2_max estimation

Our approach for estimating VO_2_max from UKB CRF test HR response is illustrated in Figure 1B-E. Here we first describe a VO_2_max estimation method for HR response to steady-state exercise. We then adapt this method to the UKB CRF test by harmonising HR response features extracted from flat and ramped UKB CRF tests to those extracted from steady-state exercise.

#### Conceptual framework

VO_2_max can be estimated from HR response to exercise at steady-state WR increments using linear extrapolation of the submaximal HR-to-WR relationship ^28,29^. For this approach, an individual exercises at two or more submaximal WR increments while HR is recorded. The steady-state HR response at each test increment is then regressed against WR to establish a line-of-best fit for the observed HR-to-WR relationship (W ⋅ bpm^-1^). This relationship can be represented as:

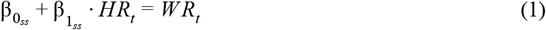

where *WR*_*t*_ and *HR*_*t*_ are paired measurements at several test increments, 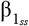 is the linear regression slope representing the steady-state HR-to-WR relationship, and 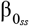 is the intercept of that regression. The regression line is extrapolated to age-predicted maximal HR (HRmax) ^30^ to estimate the WR that would be achieved if the exercise test was completed to exhaustion (i.e. the respiratory compensation point; RCP ^31,32^). VO_2_max is then estimated by converting the extrapolated WR value to net VO_2_ using a caloric equivalent of oxygen and adding an estimate of resting VO_2_ plus the VO_2_ required for unloaded cycling ^11^.

The HR-to-WR linear extrapolation approach presents challenges when applied to ramped exercise HR response. Assuming HR and VO_2_ responses are linearly related and after accounting for differences in onset kinetics ^33,34^, the principal methodological issues are ^32,35,36^: 1) within-participant, the VO_2_-to-WR relationship and total time delay for VO_2_ response to achieve linearity after ramped exercise onset will vary across ramped tests as a function of ramp rate; 2) The ramped VO_2_-to-WR relationship decreases asymptotically with ramp rate and, as ramp rate approaches zero, becomes similar to values determined from steady-state exercise; 3) the VO_2_-to-WR relationship has high test-retest variability; and 4) the VO_2_-to-WR relationship diverges from linearity above RCP. Thus, the HR-to-WR linear extrapolation approach will induce VO_2_max overestimation bias as a function of ramp rate, demonstrate low test-retest reliability, and have poor precision if the WR computed at age-predicted HRmax is greater than the WR corresponding to the RCP.

#### Multilevel modeling framework

We addressed these methodological issues by constructing a multilevel modeling framework that estimates a participant’s steady-state HR-to-WR relationship using features extracted from HR responses across UKB CRF test protocols. Our modeling framework has three levels; the first equates WR computed from steady-state HR response (Equation 1) with WR computed from regression coefficients that vary between and within individual participants (i.e. dynamic regression coefficients). Within every *i*^*th*^ individual participant, each having completed a set of *p* exercise protocols:

Level-1 (base-level equating steady-state test HR response with UKB CRF flat, low ramped, and high ramped HR response):

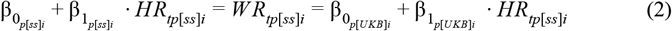

where: 1) 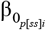 and 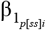 are linear regression coefficients estimated from the steady-state protocol (*p[ss]*); 2) *HR*_*tp*[*ss*]*i*_ is a sequence of *t* simulated steady-state HR values, equally spaced and spanning the submaximal intensity range; 3) *WR*_*tp*[*ss*]*i*_ is a sequence of *t* steady-state WR values computed with 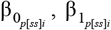, and *HR*_*tp*[*ss*]*i*_ thus, a matrix representation of the line defined by Equation 1); and 4) 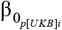 and 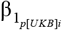 are dynamic regression coefficients that, while unique to each UKB protocol (*p[UKB]*) and individual, converge to the values of 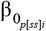 and 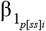 by their linkage with *WR*_*tp*[*ss*]*i*_. 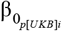 and 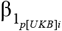 are estimated at the second level of the modeling framework using combinations of HR-response and protocol-based features:

Level-2 (HR-response and protocol features extracted from flat and ramped UKB CRF tests):

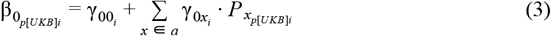

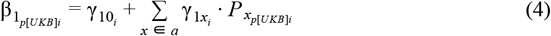

where: 1) 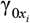 and 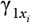 are sets of *a* fixed regression coefficients for HR-response and protocol-level features 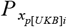; and 2) 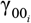 and 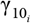 are the mean intercept and slope for the *i*^*th*^ individual participant. 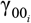 and 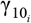 are estimated at the third level of the modeling framework using pretest participant characteristics:

Level-3 (pretest participant characteristics):

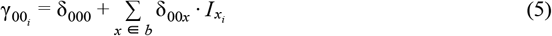

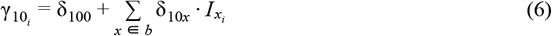

where: 1) δ_00*x*_ and δ_10*x*_ are sets of *b* fixed regression coefficients for participant characteristics 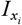; and 2) δ_000_ and δ_100_ are the model-invariant intercept and slope. 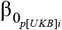 and 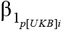 can be estimated using different sets of HR-response and protocol features 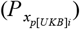 and sets of participant characteristics 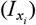. We leveraged this adaptability to derive five nested WR estimation equations (notated as M1-M5; see Supplemental Table 3), each using different combinations of feature sets, so that our approach was robust to data quality issues encountered when analysing HR response data. Additional details regarding model optimisation and the extraction of feature sets included in 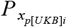 and *I* 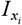 are provided in Supplemental Methods.

#### Application of estimation model

In validation study and UKB participants, VO_2_max was estimated using the set of nested WR estimation equations by extrapolating the linear fit defined 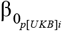 and 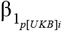 to age-predicted HRmax ^30^ and converting the resulting WR value to VO_2_max using the American College of Sports Medicine metabolic equation for cycle ergometry ^10^. We also estimated WR and VO_2_max values using a simple linear regression approach ^20^ and a similar approach for steady-state tests ^16^ (see Supplemental Methods) and compared their validity with the set of nested WR estimation equations.

#### Agreement analyses

We used Bland-Altman analysis to quantify agreement between estimated WR and VO_2_max values with those directly measured during the maximal ramp test. Correlations between estimated and directly measured values were quantified using Pearson’s *r* and Spearman’s *rho*. Estimation model precision was expressed as the root mean square error (RMSE) between estimated and directly measured values. One-sample t-tests were performed to determine whether mean biases were statistically significantly different from zero mean bias. ANOVA repeated measures were used to test differences between estimated and directly measured values across estimation models.

#### Short-term test-retest reliability

To assess short-term test-retest reliability, a subsample of 87 validation study participants completed a second UKB CRF test within 2 weeks after main testing, identical to either the low or high ramped test at the main visit. Estimated VO_2_max values from first and second tests were compared using agreement analysis.

### Estimation of VO_2_max and health associations in the UKB cohort

#### UKB participants

The UKB is a prospective cohort study of 502,625 older adults. Baseline data collection was conducted between 2006 and 2010 where a variety of physical measurements, biological samples, and health questionnaires were administered; repeat-measures visits were conducted between 2012 and 2013. The UKB CRF test was offered approximately 100,000 times (last 79,209 participants from baseline and 20,218 from the repeat-measures visit). Supplemental Figure 2 describes criteria used to assign WR estimation equations derived from the multilevel modelling framework; Supplemental Figure 3 demonstrates the results of this process. VO_2_max values were estimated as described, previously; however, age-predicted HRmax was reduced by 20bpm in those taking beta-blockers ^37^.

**Figure 2.**
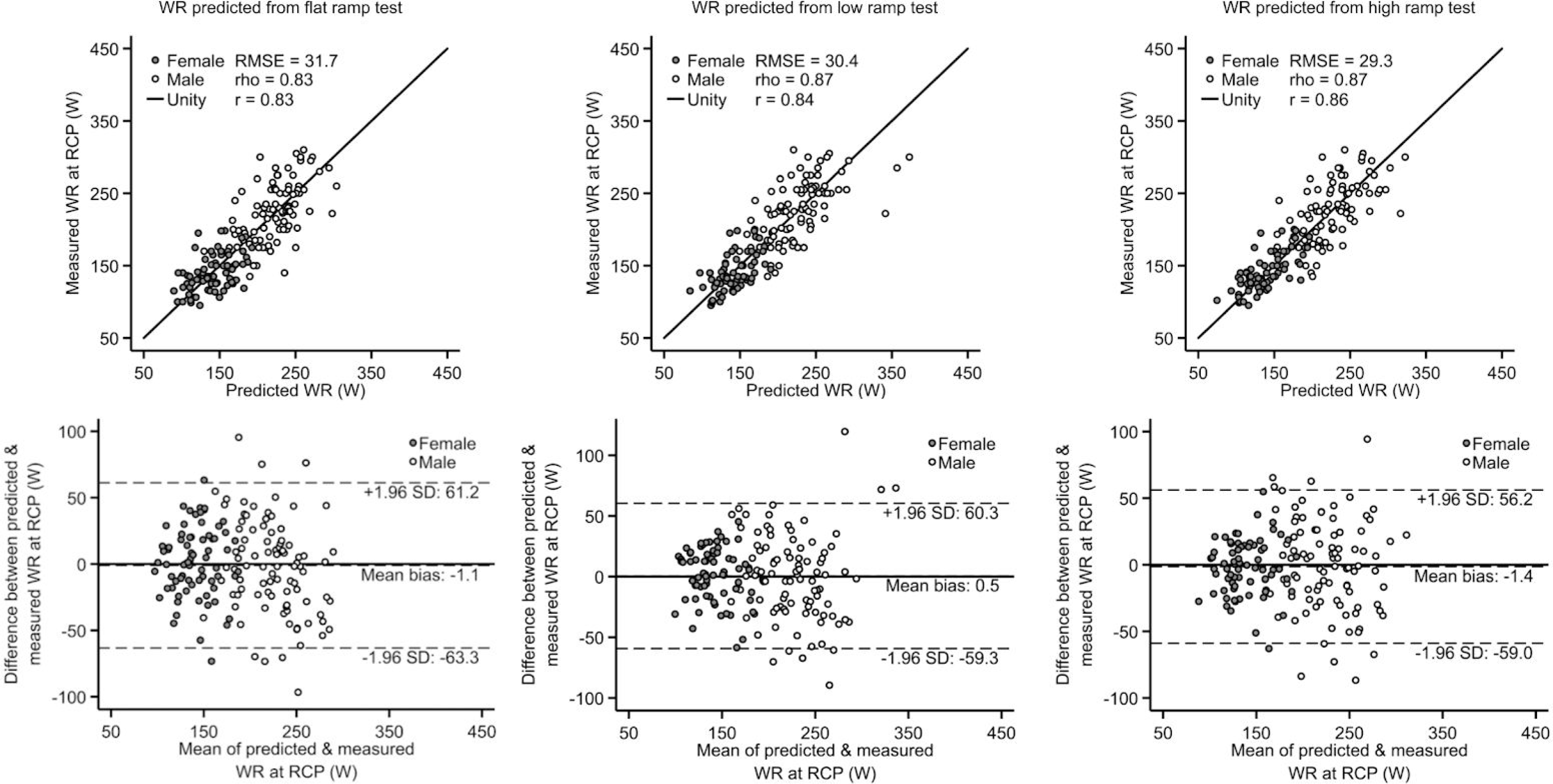
Scatterplots (top row) and Bland-Altman plots (bottom row) demonstrating agreement between work rates measured at the respiratory compensation point (RCP) and work rates estimated from flat ramp tests (left column), low ramp tests (middle column), and high ramp tests (right column) using the most comprehensive prediction equation from the multilevel modelling framework (M1 for ramp tests; M4 for flat test). r: Pearson’s correlation coefficient, rho: Spearman’s rank correlation coefficient. RMSE: Root-mean-square error.

**Figure 3.**
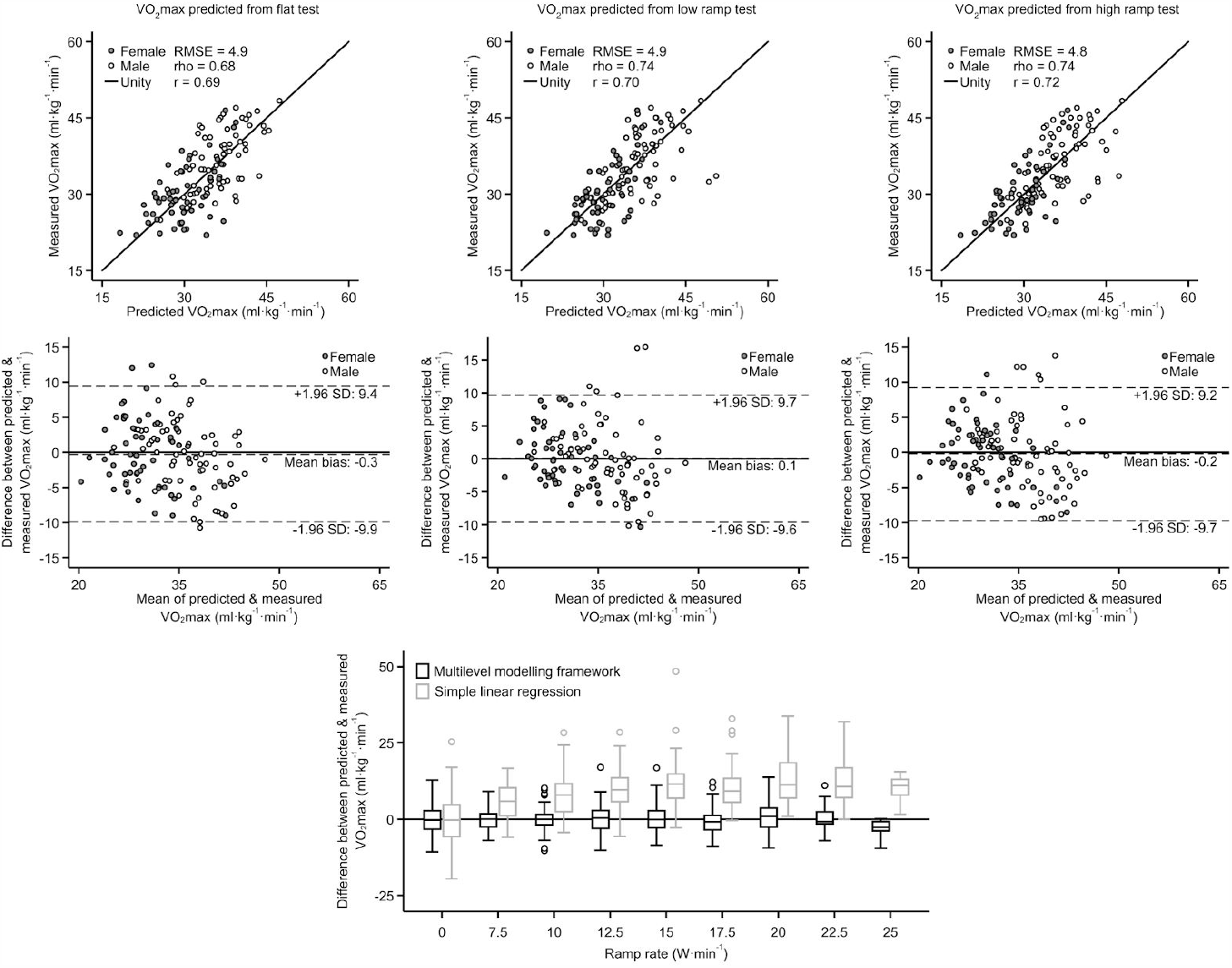
Scatterplots (top row) and Bland-Altman plots (second row) demonstrating agreement between directly measured VO_2_max and VO_2_max estimated from flat tests (left column), low ramp tests (middle column), and high ramp tests (right column) using the most comprehensive equation from the multilevel modelling framework (M1 for ramp tests; M4 for flat test). Below these (bottom row), a box plot demonstrates agreement across all ramp rates tested using the multilevel modelling framework and the simple linear regression approach. r: Pearson’s correlation coefficient, rho: Spearman’s rank correlation coefficient. RMSE: Root-mean-square error.

#### Health characteristics across CRF levels in UKB

Health characteristics were described across age-adjusted and sex-specific CRF categories ^36^. We age-stratified the UKB cohort in half-decades as <50, 50 to 54, 55 to 59, 60 to 64, and ≥65 years, defined CRF categories by tertiles (“lower”, “middle”, and “higher”) of estimated VO_2_max levels from each age stratum, and combined CRF categories from each age stratum to form CRF categories for the entire UKB cohort. Health characteristics were compared across CRF tertiles for men and women separately.

#### Survival analyses

Cox regression with age as the underlying timescale was used to estimate linear associations between estimated VO_2_max levels (in METs; 1 MET = 3.5 ml O_2_·kg^-1^·min^-1^) and mortality and incident disease outcomes. We compared prospective associations between two VO_2_max estimation approaches: the multilevel modeling framework developed in this study and the previously described method using simple linear regression. Vital status and primary or secondary hospital episodes of UKB participants were established by linkage to national registry data obtained from the Health and Social Care Information Centre (now NHS Digital) for England and Wales and the Information Services Department (ISD) for Scotland. The censoring date for mortality outcomes was 31^st^ March 2020. Censoring dates for incident disease outcomes were 31^st^ January 2018 in England and Wales, and 30^th^ November 2016 in Scotland. International Classification of Diseases (ICD) 10^th^ edition (ICD-10) codes were used to define health outcomes (See Supplemental Materials). Models were adjusted for age, sex, body weight, ethnicity, smoking status, employment status, Townsend index of deprivation, alcohol consumption, red meat intake, medication use (beta blockers, calcium channel blockers, ACE inhibitors, diueretics, bronchodialators, lipid-lowering agents, iron deficiency anaemia treatments), hypertension, diabetes, and pre-baseline self-report and hospital episodes of heart failure, ischaemic heart disease, stroke, or cancer. Potential residual confounding by obesity was addressed in stratified analyses. Participants experiencing disease events in the first two years of follow-up were excluded (analysis specific). Nonlinear associations between estimated VO_2_max levels and each of the health outcomes were evaluated using a cubic spline regression model with three knots placed at the 25^th^, 50^th^, and 75^th^ percentiles of the VO_2_max distribution. Spline models were adjusted using all covariates listed above and with 8.0 METs chosen as a reference point for the estimation of hazard ratios.

#### Long-term test-retest reliability

To assess long-term test-retest reliability, we compared estimated VO_2_max values at baseline and the first follow-up test (n = 2877, mean follow-up time 2.8 years). The follow-up UKB CRF test protocol was re-individualised at the time of testing and therefore may have differed from the baseline protocol.

All analyses were performed in Stata/SE 15.1 (StataCorp, Texas, USA). Statistical significance was set at *p* < 0.05.

## Results

### Validation of UKB CRF test

Validation study participant characteristics are described in Table 1. We recruited 105 women (mean age: 54.3y ± 7.3) and 86 men (mean age: 55.0y ± 6.5), all of whom were included in the development of our multilevel modeling framework. Data from some participants were excluded from further analyses due to issues with the integrity of HR and VO_2_ response data from the maximal exercise test (n = 25) and for failure to achieve predefined VO_2_max threshold criteria (n = 33). Participant subsample characteristics were generally similar in each subsequent validity analysis (Supplemental Figure 4).

**Figure 4.**
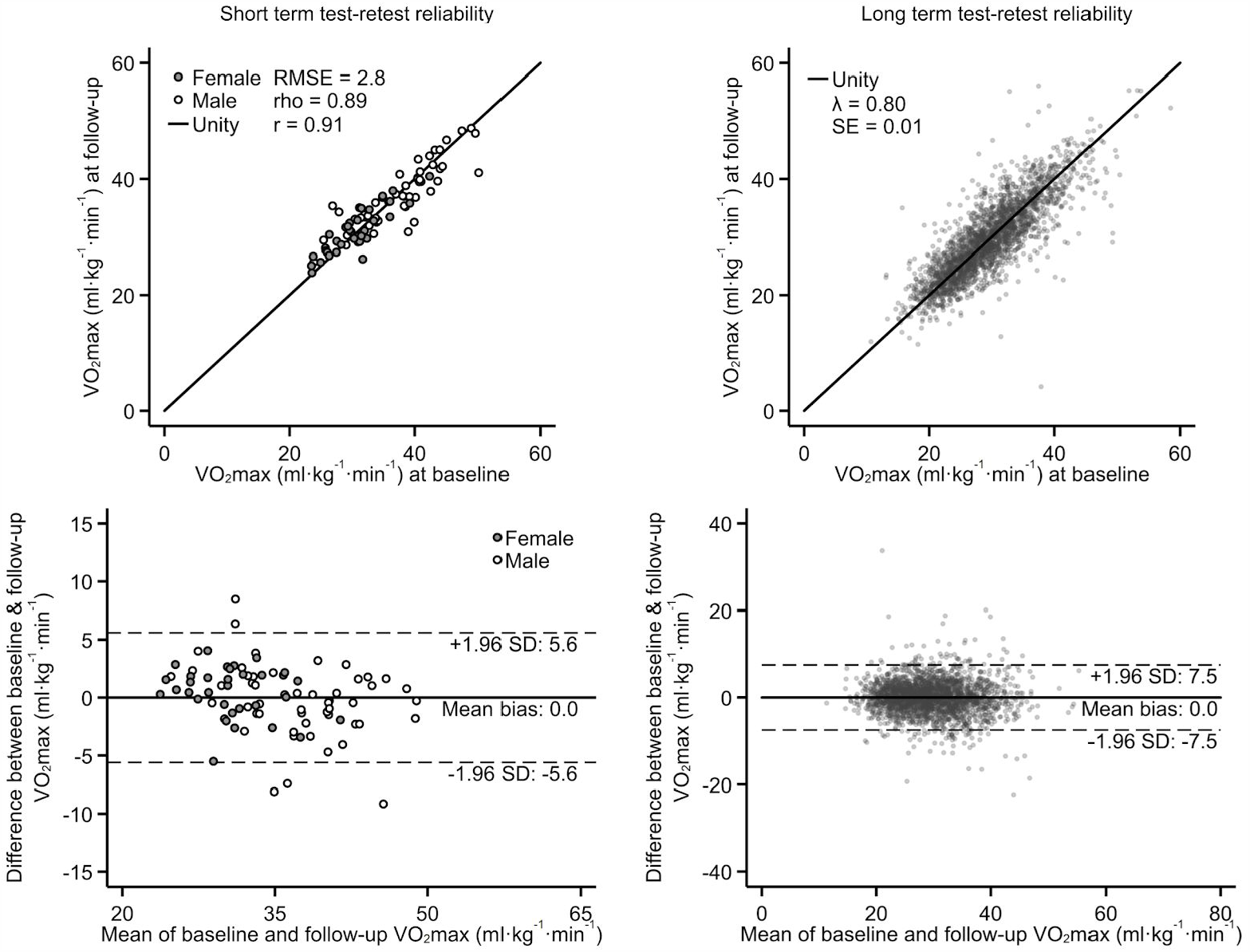
Scatterplots (top row) and Bland-Altman plots (bottom row) demonstrating short- and long-term test-retest reliability. r: Pearson’s correlation coefficient, rho: Spearman’s rank correlation coefficient. RMSE: Root-mean-square error. Lamda: Regression-dilution coefficient. SE: Standard error. Short-term reliability data are from the validation study (n = 87, follow-up ∼10 days) and long-term reliability data are from the repeat-measures substudy in UK Biobank (n = 2877, follow-up ∼2.8yrs).

The maximal WR estimated from our modelling framework (Supplemental Table 3) was compared with WR measured at the respiratory compensation point (RCP) during the maximal test. This is shown in Figure 2 for the top-level equation; levels of agreement for all subequations are shown in Supplemental Table 4. Across estimation equations (M1 through M5), estimated WR were strongly correlated to observed WR at RCP (Pearson’s *r* range: 0.81 to 0.86) with no significant mean bias in both women (Bias range: −3.7 to 3.8) and men (Bias range: −5.2 to 0.1). WR agreement did not differ between low and high ramped tests, but precision was lower for flat tests. Correlation strength was higher when WR was computed using features from ramp- and recovery-phase data (models M1 through M3) compared to using only flat-phase data (models M4 and M5), although all models were relatively precise. Estimated maximal WR did not agree with observed WR at LT (Supplemental Table 5) and at VO_2_max (Supplemental Table 6).

The maximal WR estimated from our modeling framework was converted to estimated VO_2_max and compared with VO_2_max directly measured during the maximal test as shown in Figure 3 for the top-level estimation equation; results for all subequations are shown in Supplemental Table 7. Estimated VO_2_max was correlated (Pearson’s *r* range: 0.68 to 0.74) to measured VO_2_max with no significant mean bias in both women (Bias range: −0.8 to 0.4) and men (Bias range: −0.3 to 0.3). To evaluate the internal validity of estimated VO_2_max, we compared estimated VO_2_max values from low and high ramp tests across estimation equations M1-3 and M5, as well as between flat tests across M4 and M5 (Supplemental Table 8). VO_2_max estimates from different UKB CRF test protocols were highly correlated across estimation levels (Pearson’s *r* range: 0.94 to 0.99) with low or nonsignificant bias (Bias range: −0.6 to 0.0). Estimation bias across different protocol ramp rates was also evaluated (from 0 W ⋅min^-1^ for the flat test and 7.5 to 25 W ⋅min^-1^ for the ramp tests); mean estimation bias did not differ across all ramp rates tested.

As a sensitivity analysis, we compared directly measured VO_2_max with VO_2_max values estimated using measured HRmax (Supplemental Table 9); agreement only improved marginally using measured versus age-predicted HRmax. We also tested the validity of VO_2_max estimation from a simple linear regression method (Supplemental Figure 5), which assumes no differential bias across UKB CRF tests with different ramp rates. The simple linear regression method demonstrated considerable overestimation bias and low precision when applied to ramped tests but was unbiased when applied to flat tests.

**Figure 5.**
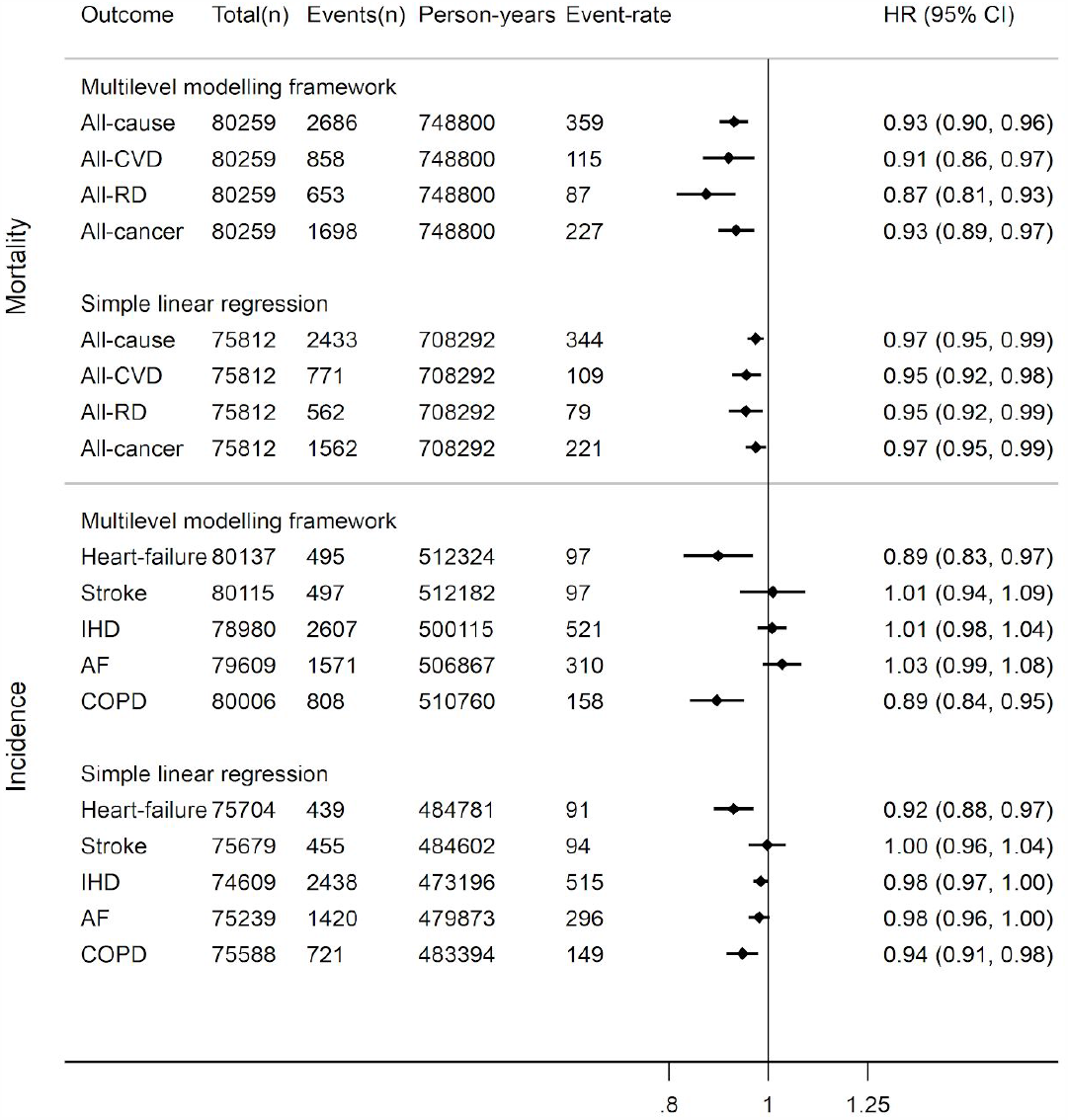
Hazard ratio (HR) and 95% confidence interval (CI) for prospective log-linear associations (Cox regression) between fatal and non-fatal outcomes in the UK Biobank with cardiorespiratory fitness in metabolic equivalents (METs, per 3.5 ml O_2_·kg^-1^·min^-1^) estimated from the multilevel modelling framework and simple linear regression methods. Event-rate per 100,000 person years. AF: atrial fibrillation; COPD: chronic obstructive pulmonary disease; CVD: cardiovascular disease; IHD: ischaemic heart disease; RD: respiratory disease. COPD incidence mostly reflects severe COPD since only ∼25% of cases end up in hospital. Cumulative mortality and incidence rates differ between fitness prediction methods owing to different inclusion criteria at the estimation level.

We evaluated the short- and long-term test-retest reliability of the UKB CRF test in validation study and UKB study participants, respectively (Figure 4). Estimated VO_2_max values from the first and second tests were highly correlated with no mean difference for test-retest within two weeks, and nearly as strong over the long-term.

### Cardiorespiratory fitness and health associations in UKB cohort

Table 2 describes UKB participant health characteristics by sex and CRF strata defined using VO_2_max values estimated from our modelling framework. Estimated VO_2_max was higher in men compared to women, and in younger versus older adults. Participants in the middle and higher CRF tertiles had better baseline measures of heart and lung function, lower body weight, and better self-perceived health than participants in the lower tertile.

In total, 2686 participants died during a median 9.9 years (interquartile range 9.7 to 10.0 years) of follow-up (749,136 person-years). After adjustment for potential confounders, every 1-MET difference in CRF was associated with approximately 7% lower all-cause mortality; associations were stronger for deaths from respiratory disease (RD), cardiovascular disease (CVD), and cancers (Figure 5), and also stronger in the obese (Supplemental Figure 6). Incidence of chronic obstructive pulmonary disease and heart-failure were more strongly associated with differences in CRF than stroke, ischaemic heart disease, atrial-fibrillation, and cancers; only the COPD association was significant and stronger in the obese (Supplemental Figure 6). Compared to associations computed using the simple linear regression method, health associations were generally stronger but estimated with more uncertainty when using VO_2_max levels estimated from our multilevel modeling framework.

**Figure 6.**
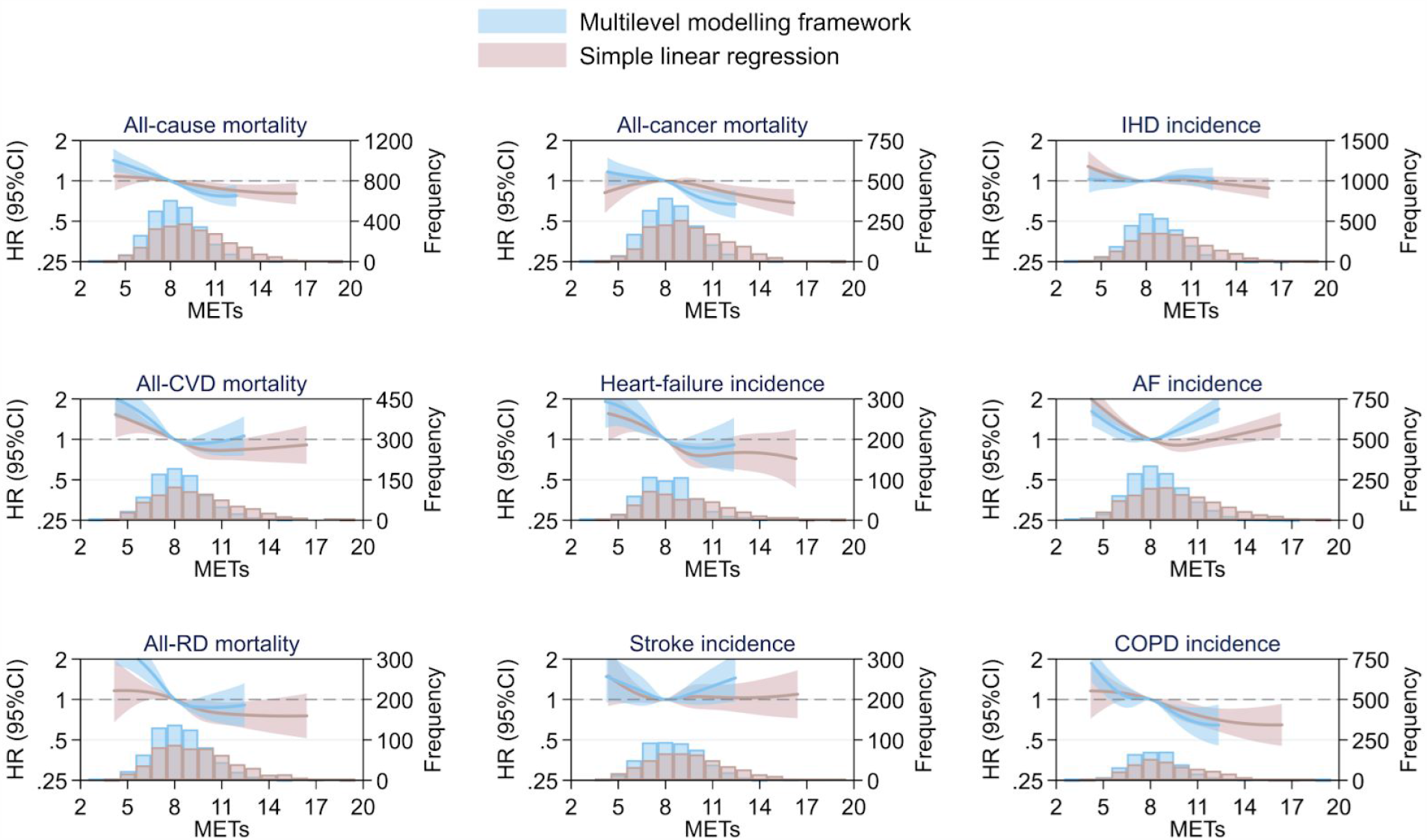
Hazard ratio (HR) and 95% confidence interval (CI) for nonlinear associations (cubic splines, Cox regression) between fatal and non-fatal outcomes in the UK Biobank with cardiorespiratory fitness in metabolic equivalents (METs, per 3.5 ml O_2_·kg^-1^·min^-1^) estimated from the multilevel modelling framework and simple linear regression. Hazard ratios were computed relative to a fitness reference point of 8.0 METs. AF: atrial fibrillation; COPD: chronic obstructive pulmonary disease; CVD: cardiovascular disease; IHD: ischaemic heart disease; RD: respiratory disease. Cumulative mortality and incidence counts (superimposed histograms) differ between fitness estimation methods owing to different inclusion criteria at the estimation level.

Dose-response relationships between CRF levels (in METs) estimated from our multilevel model and mortality as well as incident disease outcomes are shown in Figure 6, with obesity-stratified results in Supplemental Figure 7. CRF was inversely associated with mortality from all causes, CVD mortality, RD mortality, and cancer mortality for the range of 3-11 METs. The shape of CRF dose-response relationships varied considerably across incident disease outcomes. In the range of 3-8 METs, CRF was inversely associated with incidence of CVD from all causes, IHD, heart-failure, AF, stroke, RD, and COPD; disease associations flattened (IHD, heart failure, RD, COPD) or became positive (CVD, AF, stroke) above 8 METs. The association between CRF and incidence of cancers demonstrated an inverted-U relationship. Differences between these associations and those observed using the simple linear regression to estimate CRF were most evident at the tails of the distributions. Supplemental Figures 8 and 9 demonstrate linear and nonlinear survival sensitivity analyses for both CRF estimation methods but in the same analytical sample. These sensitivity analyses were further restricted in Supplemental Figures 10 and 11 by limiting the estimation of CRF to model M5 for the multilevel modeling framework.

## Discussion

In the largest and most inclusive population-based study of fitness known to date, we present a valid and reliable method for estimating VO_2_max and demonstrate its utility in characterising associations with disease endpoints in UK adults. Our method uses individual HR response to a risk-stratified and individualised ramped exercise protocol (i.e. the UKB CRF test) to harmonise CRF estimation using a unifying modeling framework anchored in steady-state exercise response. We show in an independent validation study that maximal WR estimated from our method corresponds to an individual’s respiratory compensation point and, when converted to VO_2_max, agrees with directly measured VO_2_max across different UKB CRF test protocols. Having resolved the validity issues of differential bias across test protocols and absolute agreement with directly measured VO_2_max, we characterise dose-response relationships between estimated fitness levels and all-cause and cause-specific mortality and morbidity in UK adults, demonstrating that our novel approach yields relationships which are on average twice as strong as those reported using non-validated approaches.

CRF-health associations reported in this study are in agreement with numerous other populations-based studies demonstrating the protective effects of CRF on all-cause and cause-specific mortality and morbidity ^1–3^. We did not find associations for fatal and non-fatal incidence rates of aggregated CVD, ischemic heart disease, and respiratory disease.

In agreement with previous studies, associations were J-shaped for stroke ^38^ and U-shaped for atrial fibrillation ^39^; a study with more follow-up time and incident events, however, reported inverse relationships between CRF and atrial fibrillation ^40^. Our study found an inverse relationship between CRF and all-cancer mortality, in agreement with several previous studies ^41,42^. Additional follow-up time is warranted to investigate CRF associations with site-specific cancers in UKB participants.

The primary strength of our approach is that by conducting separate validation work, we were able to maximise the validity of estimated VO_2_max from the UKB CRF test by: 1) utilising HR response features across all test phases, thereby increasing the proportion of data used to infer the latent HR-to-WR relationship; 2) incorporating resilience in HR response feature dependency; and 3) anchoring the inferential modeling of those features - which can vary with protocol ramp rate - to the more invariant HR-to-WR relationship as estimated from steady-state testing. For these reasons, it obtains results that diverge considerably from those used in previous attempts at describing CRF in the UKB cohort. In a previous publication from our group ^16^, we estimated CRF by using simple linear regression of all recorded heart rate data during the test to estimate the HR-to-WR relationship. Results from the present study demonstrate that this approach overestimates VO_2_max differentially by ramp rate, thus limiting the ability to validly compare VO_2_max estimates from different UKB CRF tests. Our novel multilevel modeling approach, afforded by additional validation data, demonstrates stronger associations with all-cause and cause-specific mortality and morbidity compared to non-validated methods. Furthermore, given the adaptability of our multilevel modeling framework to missing or low-quality test data, we were able to include more participants in our CRF-health association analysis. Increased uncertainty around CRF-health association estimates using CRF predicted from the multilevel modelling framework are likely more accurate than those predicted with simple linear regression.

Other approaches also use simple linear regression, but establish the HR-to-WR relationship by relating resting HR to only a single measurement of HR during the test ^20,43,44,22,18,21,45,46,17^. HR measurement noise will greatly decrease precision in this approach, and the CRF estimates are still subject to bias, the extent of which may differ by protocol. Another reported approach ^19,47–49^ is to use the maximally achieved WR to infer CRF, which simply reflects the protocol that participants were assigned according to particpant age, sex, resting heart rate, and exterional chest pain risk. As the protocol was risk-stratified, prospective associations of such an exposure measure with heart disease endpoints does validate the risk stratification, but it is not possible to interpret this as an association with CRF. It is not immediately clear how conclusions reached in these previous reports might change if reexamined using the VO_2_max estimation approach developed in this study.

Our approach also has implications for exercise prescription in clinical environments, in that we have demonstrated that it is safe to test a wide range of individuals, including some of those who would normally be contraindicated for exercise. Such individuals would be prescribed a less strenuous test which does provide less information about his or her physiological state compared to a more strenuous test but the approach we outline here for interpreting such test results yields unbiased estimates of fitness. This may address a well-recognised limitation of exercise testing ^8^.

This study has several limitations. The validation study did not directly evaluate the validity of UKB CRF test protocols with ramp rates at 2.5 and 5.0 W ⋅ min^-1^. Therefore, validity of these specific ramp rates were not directly assessed but agreement for ramp rates above and below were unbiased. It is possible that our equation selection within our inference framework (Supplemental Figure 2 and 3) may differentially influence downstream CRF-health association analyses. Lastly, we examined non-fatal health outcomes using only hospitalisation data which does not necessarily capture all disease events in a given category.

## Conclusions

We have demonstrated the absolute validity, internal consistency, and test-retest reliability of a novel VO_2_max estimation method for the UKB CRF test. Our approach uses a generalised modeling framework that bridges the methodological gap between steady-state and ramped incremental exercise, addressing a persistent problem in exercise physiology and prescription. CRF estimated in this way is more strongly associated with mortality and other disease endpoints than previous methodology.

## Data Availability

The data analysed during the current study are not publicly available because we have not obtained consent for public data sharing from the study participants.

## Acknowledgements

We thank the participants of both the validation study and the UK Biobank study for their time. We are also grateful to all members of the MRC Epidemiology Unit functional groups, including field epidemiology, physical activity technical team, IT and data management for their contribution to the validation study, as well as the UK Biobank principal investigators and study teams. We thank Youngwon Kim for early discussions around the design of the validation study and Thomas White for implementation of the heart rate detection algorithms from Physionet. The work included in the present analyses use UK Biobank application #408.

This work was funded by the UK Medical Research Council (MC_UU_12015/3) and the NIHR Biomedical Research Centre in Cambridge (IS-BRC-1215-20014). UK Biobank is acknowledged for contributing to the costs of the fieldwork. The funders had no role in the design, conduct, analysis, and decision to publish results from this study.

## SUPPLEMENTAL MATERIAL

### Supplemental Methods

#### UKB CRF test description

The UKB CRF test protocol design and individualisation process are described in detail by the most recent test manual. Briefly, participants were categorised into separate risk levels according to questions adapted from the Rose Angina questionnaire. Participants with “minimal” and “small” risk completed an individualised ramp test, those with “medium” risk completed a flat test, and those with “high” risk did not complete an exercise test. Ramped tests began with a 2-minute flat-phase at a single WR (30W for females, 40W for males) followed by a 4-minute ramp-phase where WR increased continuously to a pre-specified target WR. The target WR was calculated as a risk-adjusted percentage (50% for those with “minimal” risk, 35% for “small” risk) of the maximal WR predicted from an equation derived from cycle ergometer test data collected in the Danish Health Examination Survey 2007-2008. The computed value for target WR was combined with participant sex (“F” for female, “M” for male) to notate different exercise protocols. For example, a male participant with “minimal” risk and predicted WR at VO_2_max of ∼200W would have a target work rate of 100W and be individualised to UKBB protocol “M100”. Flat tests consisted of a single 6-minute flat-phase. Participants cycled at a 60-rpm cadence while WR and HR were monitored. All tests ended with a 1-minute recovery-phase where participants sat quietly and motionless on the cycle.

#### HR response feature extraction and modeling framework

##### Feature extraction from UKBB CRF test phases

To extract features from WR and HR response data, we applied different analysis techniques to phases within each flat, ramp, and steady-state test. Data were denoted as the *t*^th^ observation in the *p*^th^ exercise protocol from the *i*^th^ individual participant. For ramp phase data (denoted *p[UKB]*), we used a simple linear regression model to describe the relationship between instantaneous WR and HR under ramped conditions:

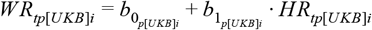

where 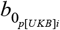 and 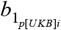 are intercept and slope parameters. HR dynamics during the recovery-phase were modelled using an exponential decay function:

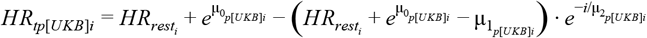

where 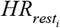 is resting HR for participant *i*. Recovery models were solved at *t* = 0s and 45s to estimate HR values at the start of recovery 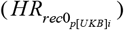 and at 45s post-recovery 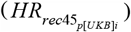. Recovery HR dynamics were also characterised using a quadratic model for comparative purposes. Flat-phase data were analysed by computing the median HR value over the last minute of the test 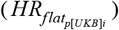. For steady-state test data (denoted as *p*[*ss*]) we used a simple linear regression model to describe the relationship between WR and HR under steady-state conditions:

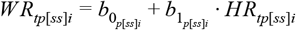

To account for delay in the achievement of a steady-state HR at each WR increment, only HR and WR data from the last minute of each increment were used to estimate 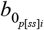 and 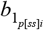.

##### Estimation of coefficients for work rate prediction models

In a two-stage procedure, we used features extracted from WR and HR response data to estimate coefficients for a WR prediction model and several nested submodels. In the first stage, intercept and slope parameters estimated from each *i*^th^ participant’s steady-state test 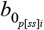 and 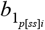 were used to estimate simulated WR values that would be achieved at a set of simulated steady-state HR values *HR*_*tp*[*sim*]*i*_:

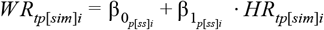

where,

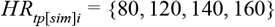

Thus, *WR*_*tp*[*sim*]*i*_ defines a set of simulated WR values achieved under steady-state test conditions for the *i*^th^ participant.

In the second stage, we combined ramp-phase linear regression parameters 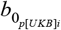 and 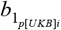, HR recovery values (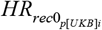 and 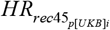), and flat-phase median HR values (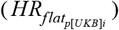) with test ramp rate (*RR*_*p[UKB]i*_), participant resting HR (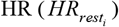) and sex (*Sex*_*i*_) to construct a multilevel modelling framework for predicting each participant’s set of simulated steady-state WR values *WR*_*tp*[*sim*]*i*_:

Level 1 (base-level equating steady-state test HR response with UKB CRF flat, low ramped, and high ramped HR response):

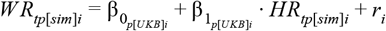

Level 2 (HR-response and protocol features extracted from flat and ramped UKB CRF tests):

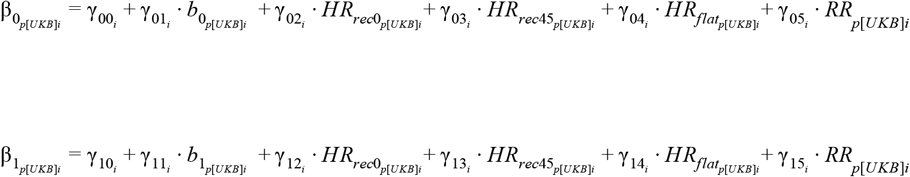

Level 3 (pretest participant characteristics):

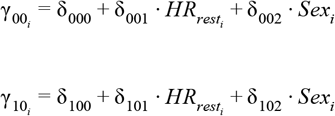

where *r*_*i*_ is a random intercept to control for clustering of observations within participants. The resulting expression for each individual will reduce to a linear equation as specified at Level 1, with dynamic test features informing the intercept and slope at Level 2, and pre-test parameters parallel-shifting these at Level 3.

We leveraged the modular structure of the multilevel modelling framework to derive a top-level work rate prediction equation and several lower-level subequations. Coefficients for the top-level equation were estimated using all derived features. Then, features were removed in a stepwise fashion to estimate coefficients for each subequation. Features were also removed to maximise explained variance with the fewest degrees of freedom. Five prediction equations, notated as M1-M5, were estimated.

##### Prediction of VO_2_max using nested prediction model

To predict VO_2_max, work rate values were estimated using the top-level work rate model and each submodel by substituting *HR*_*i*[*ss*]_ with age-predicted maximal heart rate:

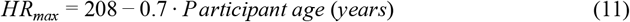

Then, estimated work rate values were converted to VO_2_ values using the American College of Sports Medicine metabolic equation for cycle ergometry:

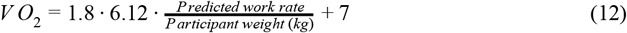

##### Prediction of VO_2_max using alternative methods

VO_2_max values were also estimated using two alternative methods. The first method, a simple linear regression approach, was applied to “low” and “high” ramp tests completed by participants:

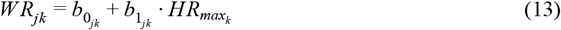

where 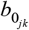 and 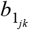 are intercept and slope parameters described previously in the ramp phase test analysis and 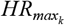 is age-predicted maximal heart rate. The second method was applied to flat tests:

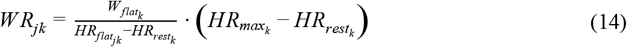

where 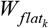 is the test steady-state work rate (30W for females; 40W for males). Work rate values were converted to predicted VO_2_max values using the ACSM metabolic equation for cycle ergometry (Equation 12).

#### VO _2_ max test analyses

VO_2_max was measured as the average of the two highest VO_2_ measurements in the last forty-five seconds of the ramped maximal exercise test. We also measured associations between work rate values computed from the multilevel modeling framework and work rates measured at several physiological events during the test (Supplemental Figure 1). Work rate values were measured at VO_2_max (i.e. maximal work rate achieved on the test, WR_max_), at the lactate threshold (LT), and at the respiratory compensation point (RCP). The work rate at LT was measured at the point when both ventilatory equivalent of oxygen (V_E_ / VO_2_) and end-tidal pressure of oxygen (P_ET_O_2_) increased with no increase in ventilatory equivalent of carbon dioxide (V_E_ / VCO_2_). The work rate at RCP was measured at the point when both V_E_ / VO_2_ and V_E_ / VCO_2_ increased and end-tidal pressure of carbon dioxide (P_ET_CO_2_) decreased. Directly measured work rates were determined visually by three independent and blinded investigators; the median value among investigators was considered the final value.

#### ICD-10 codes for non-fatal and fatal health outcomes

Non-fatal outcomes were hospital episodes of heart failure (ICD-10 codes I50, I110, I130, I132), stroke (ICD-10 codes I60-166), ischaemic heart disease (ICD-10 codes I20-I25), atrial fibrillation (ICD-10 code I48), all cardiovascular disease (CVD; ICD-10 codes I5-I9, I10-I89), chronic obstructive pulmonary disease (ICD-10 code J44), all respiratory disease (ICD-10 codes J00-J99), and all cancer (ICD-10 codes C00-99 and D00-D49). Fatal outcomes were all-cause mortality, CVD mortality, respiratory disease mortality, and cancer mortality.

## Supplemental Figures

**Supplementary Figure 1.**
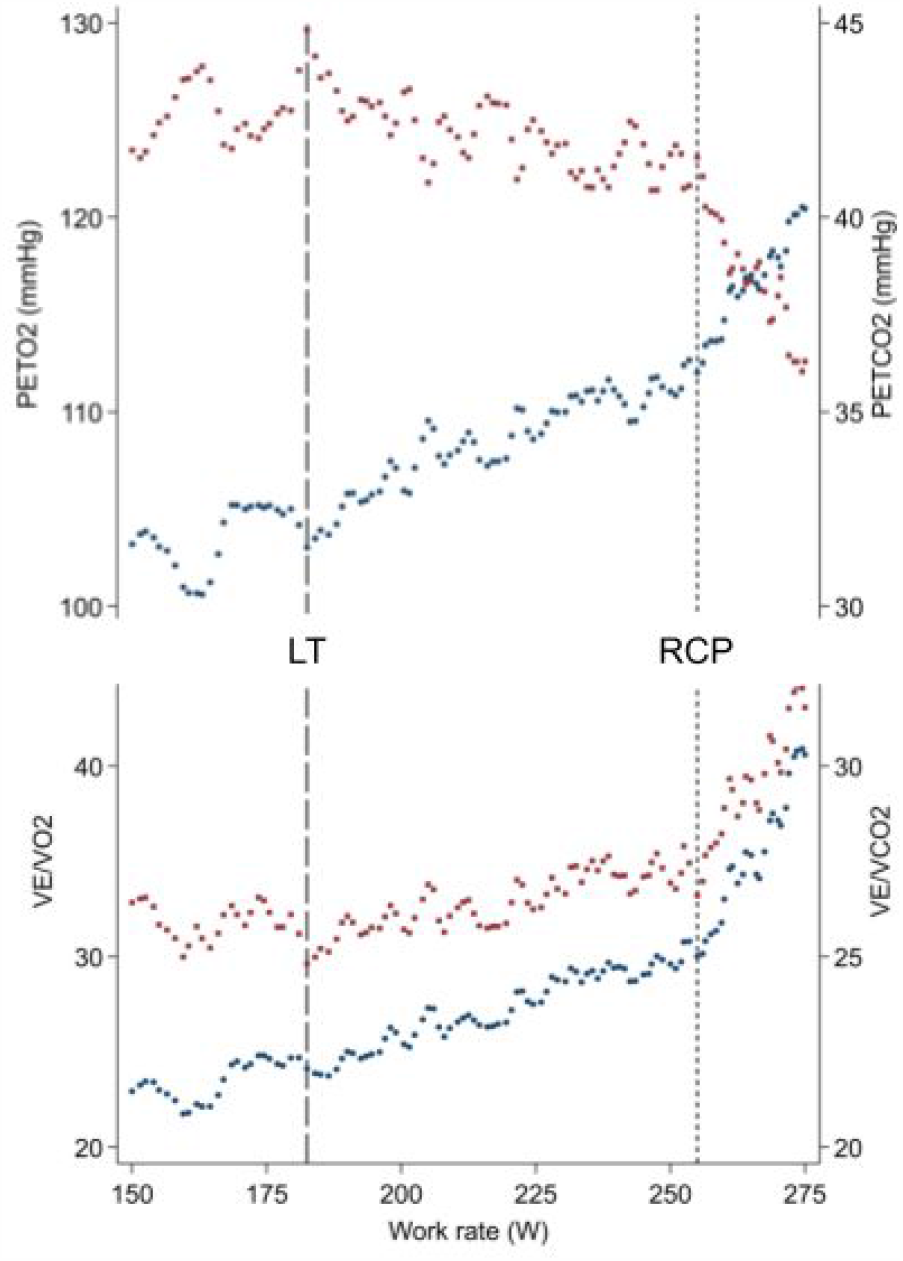
Exemplar respiratory exchange data from the ramped maximal exercise test. Work rates corresponding to the lactate threshold (LT) and respiratory compensation point (RCP) were determined by visual inspection of data representing the ventilatory equivalent of oxygen (VE / VO2, lower panel blue dot plot), ventilatory equivalent of carbon dioxide (VE / VCO2, lower panel red dot plot), end-tidal pressure of oxygen (PETO2, upper panel blue dot plot), and end-tidal pressure of carbon dioxide (PETCO2, upper panel red dot plot).

**Supplementary Figure 2.**
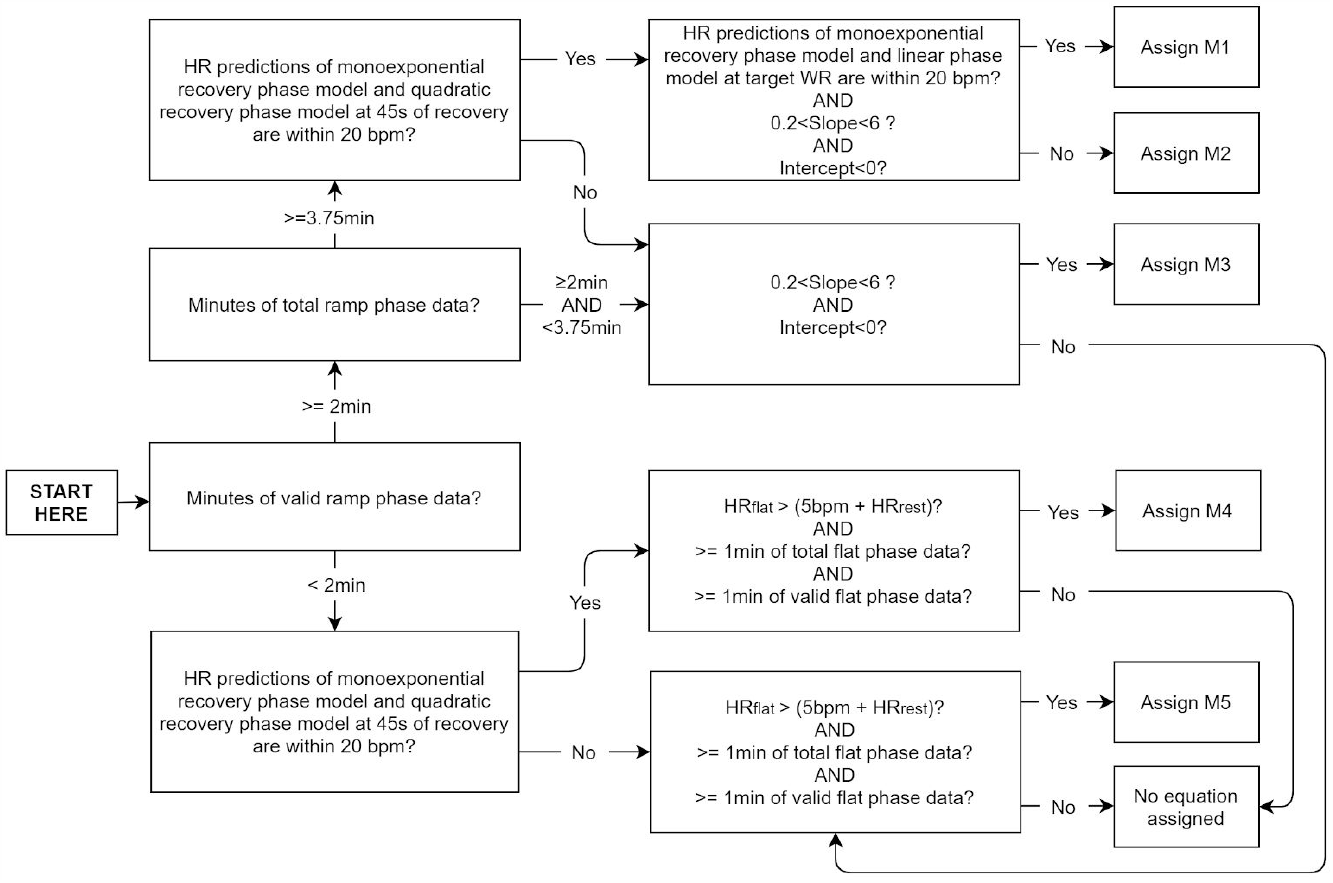
Decision diagram for the allocation of estimation equations to UKB participants. HR predictions from the ramp phase linear model were solved at the target WR of the UKB CRF test protocol. HR predictions from recovery phase models were solved at T=0s and T=45s. Slope and intercept parameters were defined using the rampe phase linear model. Recovery phase data from the flat protocol (corresponding to equation M4) is not comparable with recovery phase data from ramped protocols (corresponding to equations M1 and M3).

**Supplementary Figure 3.**
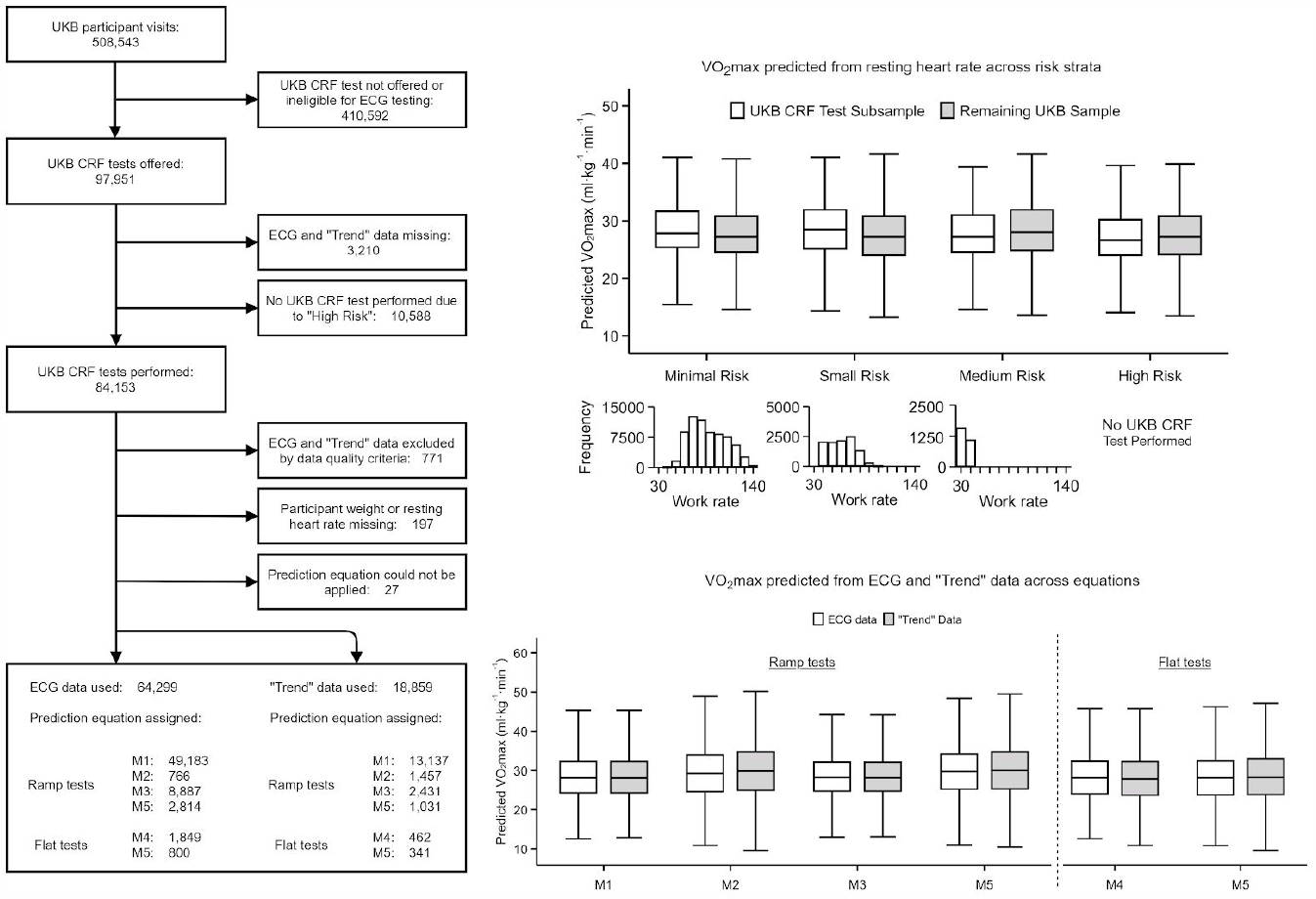
Left panel: Flow diagram showing the number of cases included and excluded in the UKB cohort analysis, as well as the allocation of work rate prediction equations. M1 represents the top-level equation with M2-M5 representing subequations. HR responses for UKB participants were recorded as either raw ECG or “Trend” data. “Trend” data represents instantaneous HR values computed using a proprietary algorithm in the software used to record data (Cardiosoft); in some tests sessions, this is the only data available (no raw ECG). Top right panel: Differences between the subsample of UKB participants with and without a bike test and stratified by eligibility, using VO_2_max estimated from resting HR within the bike test sample (VO_2_max = −0.28 RHR + 6 male sex + 44, R^2^ = 0.45, RMSE = 4.9 ml O_2_/min/kg). Histograms represent frequency of target work rates for UKB CRF tests in the subsample only across risk strata. Bottom-right panel: Sensitivity analysis comparing predicted values from ECG and “Trend” data across estimation equations and within-participant, demonstrating no differences between data capture methods. ECG data were chosen preferentially over “Trend” data when both data sources were available.

**Supplementary Figure 4.**
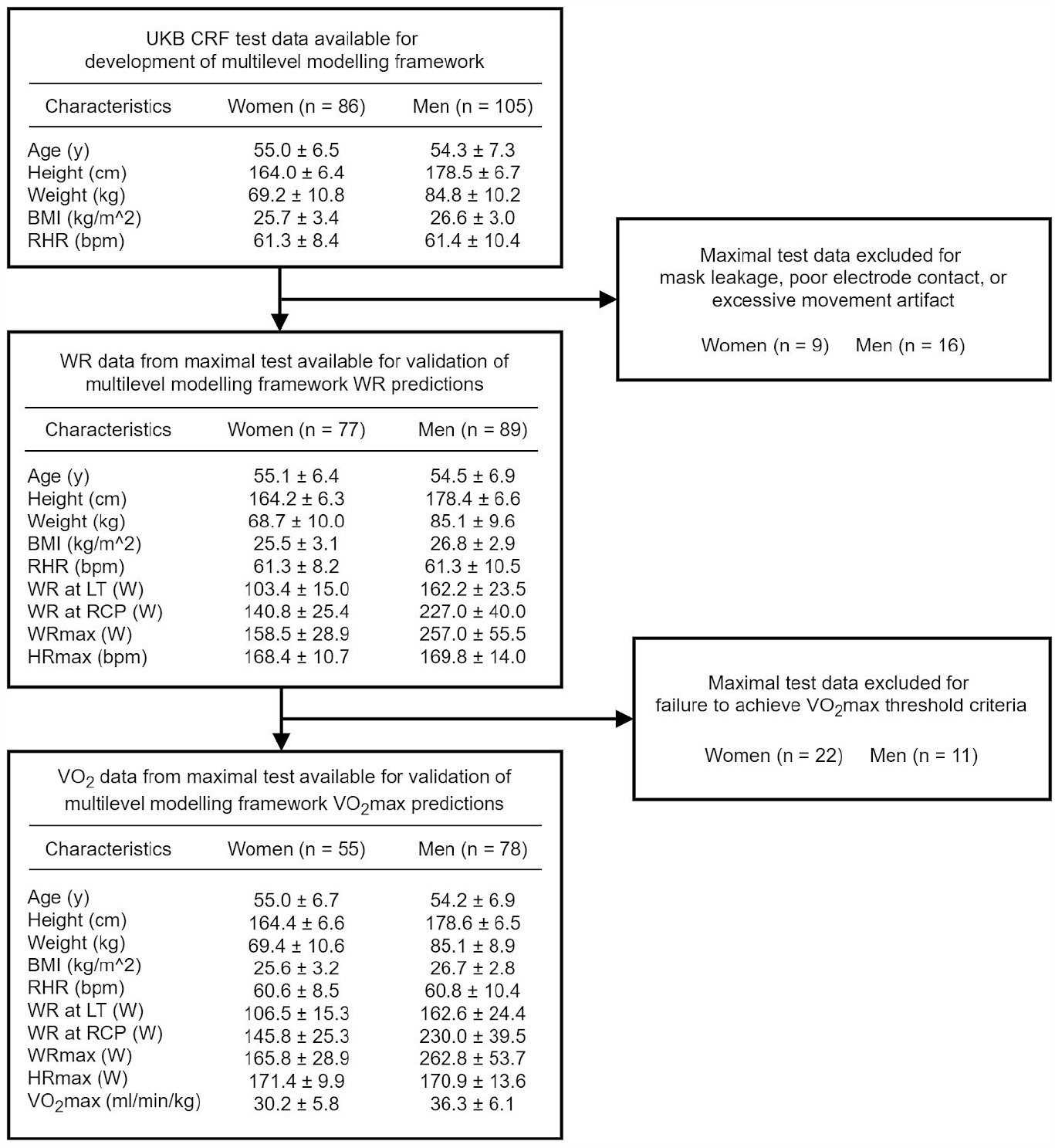
Validation study participant characteristics across each validity subanalysis. BMI: Body mass index, RHR: Resting heart rate, WR: Work rate, LT: Lactate threshold, RCP: Respiratory compensation point, WRmax: Measured maximal work rate, HRmax: Measured maximal heart rate, VO_2_max: maximal oxygen consumption

**Supplementary Figure 5.**
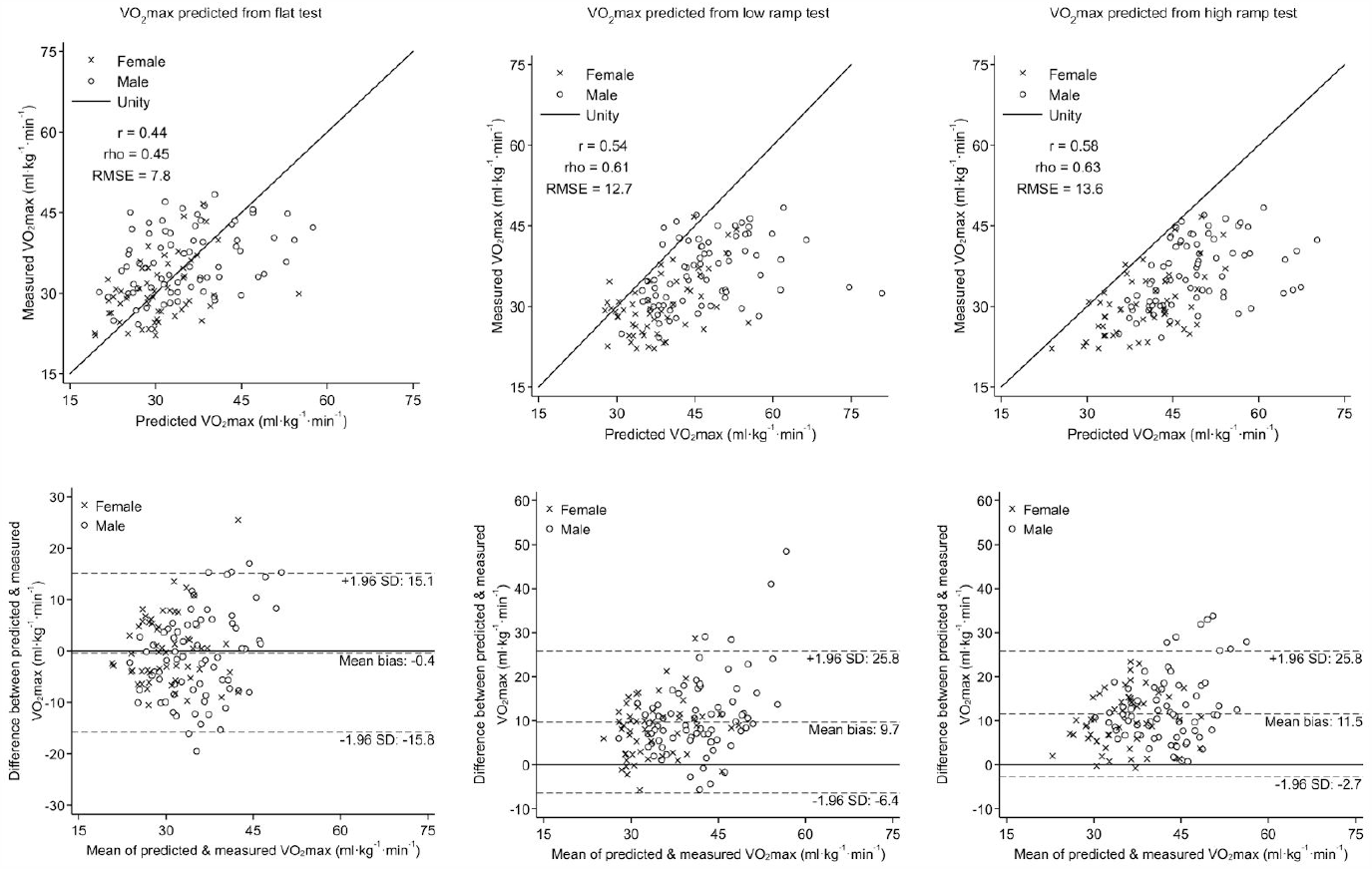
Scatterplots and Bland-Altman plots demonstrating agreement between directly measured VO_2_max and VO_2_max estimated from the flat test, low-ramp test, and high-ramp test using simple linear regression. r: Pearson’s correlation coefficient, rho: Spearman’s rank correlation coefficient. RMSE: Root-mean-square error.

**Supplementary Figure 6.**
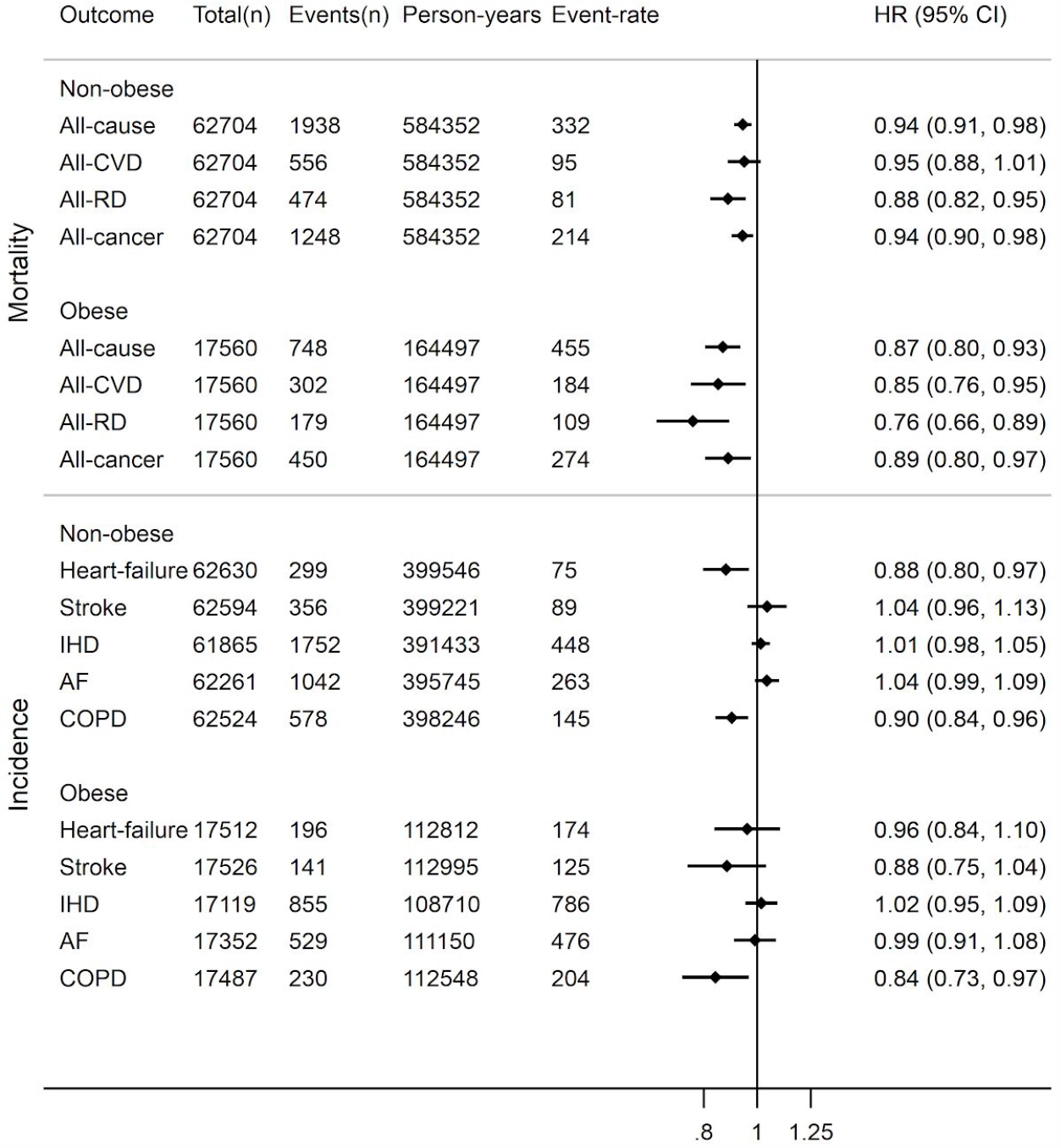
Hazard ratios (HR) and 95% confidence intervals (CI) for prospective log-linear associations between fatal and non-fatal outcomes in the UK Biobank with cardiorespiratory fitness in metabolic equivalents (METs, per 3.5 ml O_2_·kg^-1^·min^-1^), stratified by obesity status in UKB participants. Event-rate per 100,000 person years. AF - atrial fibrillation; COPD: chronic obstructive pulmonary disease; CVD: cardiovascular disease; IHD: ischaemic heart disease; RD-respiratory disease. COPD incidence mostly represents severe COPD since only ∼25% of cases end up in hospital. CRF estimates were computed using the multilevel modeling framework.

**Supplementary Figure 7.**
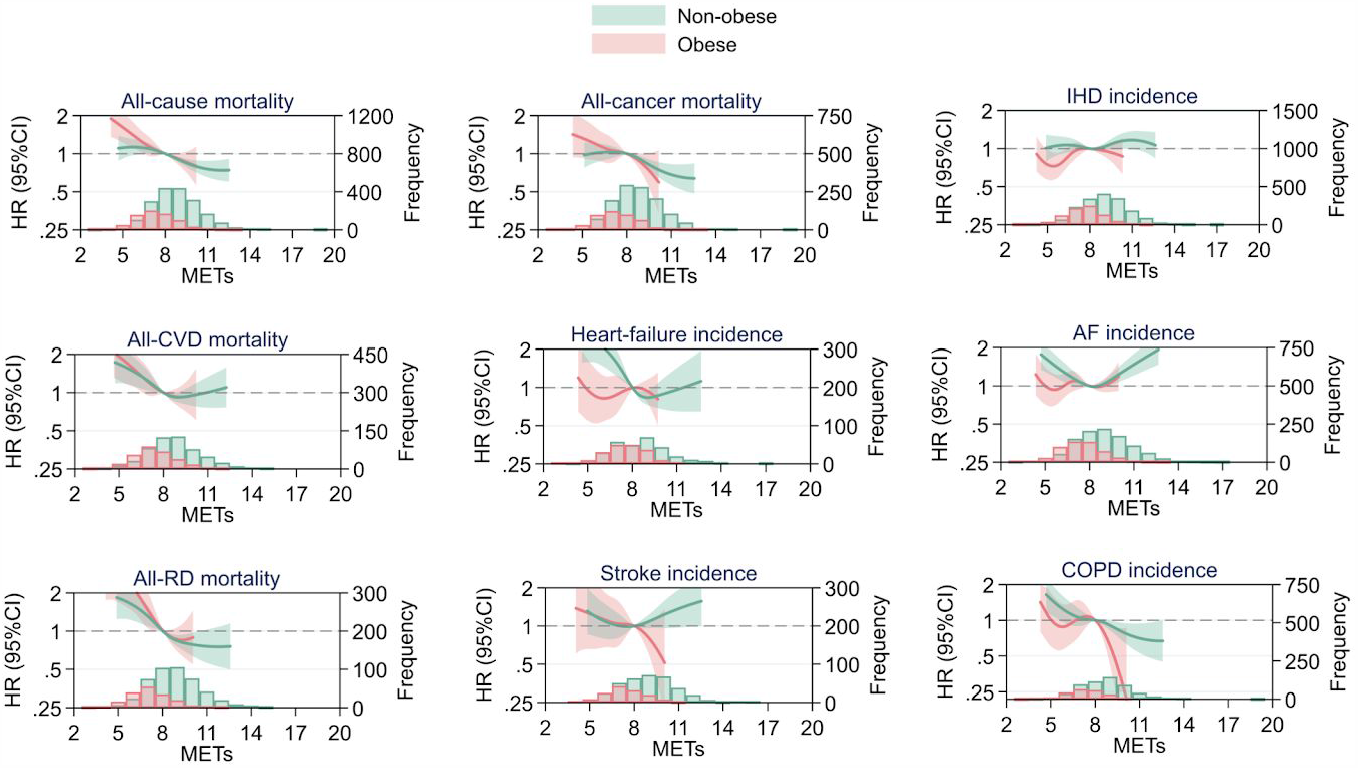
Hazard ratios (HR) and 95% confidence intervals (CI) for nonlinear (cubic spline) associations between fatal and non-fatal outcomes in the UK Biobank with cardiorespiratory fitness in metabolic equivalents (METs, per 3.5 ml O2·kg^-1^·min^-1^), stratified by obesity status in UKB participants. Hazard ratios were computed relative to a fitness reference point of 8.0 METs. AF: atrial fibrillation; COPD: chronic obstructive pulmonary disease; CVD: cardiovascular disease; IHD: ischaemic heart disease; RD: respiratory disease. CRF estimates were computed using the multilevel modeling framework.

**Supplementary Figure 8.**
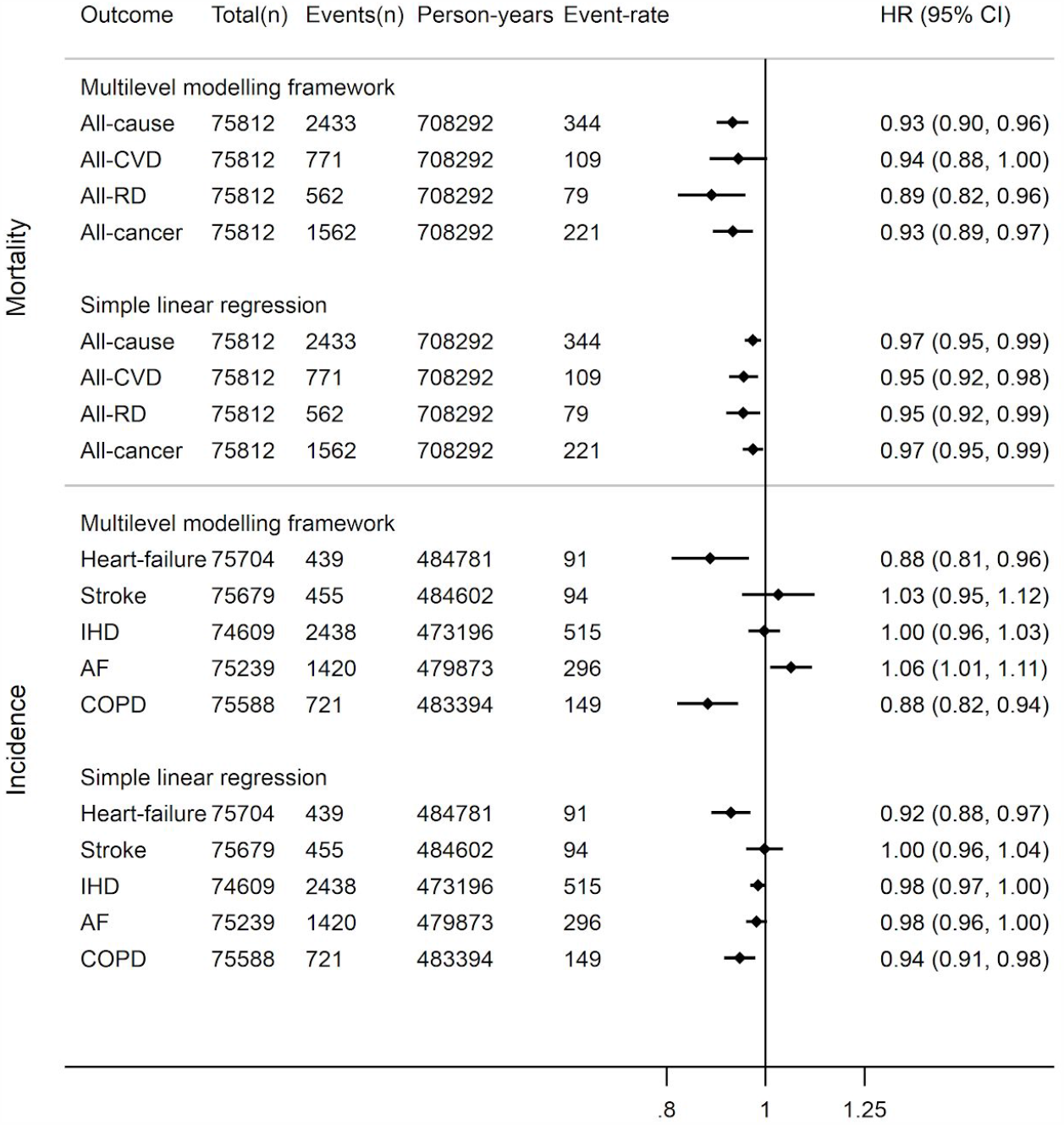
Sample-matched hazard ratios (HR) and 95% confidence intervals (CI) for prospective log-linear associations between fatal and non-fatal outcomes in the UK Biobank with cardiorespiratory fitness in metabolic equivalents (METs, per 3.5 ml O_2_·kg^-1^·min^-1^) estimated from the multilevel modelling framework and simple linear regression methods. Event rate per 100,000 person-years. AF - atrial fibrillation; COPD: chronic obstructive pulmonary disease; CVD: cardiovascular disease; IHD: ischaemic heart disease; RD-respiratory disease. COPD incidence mostly represents severe COPD since only ∼25% of cases end up in hospital. For these analyses, the analytical sample was matched between fitness estimation methods.

**Supplementary Figure 9.**
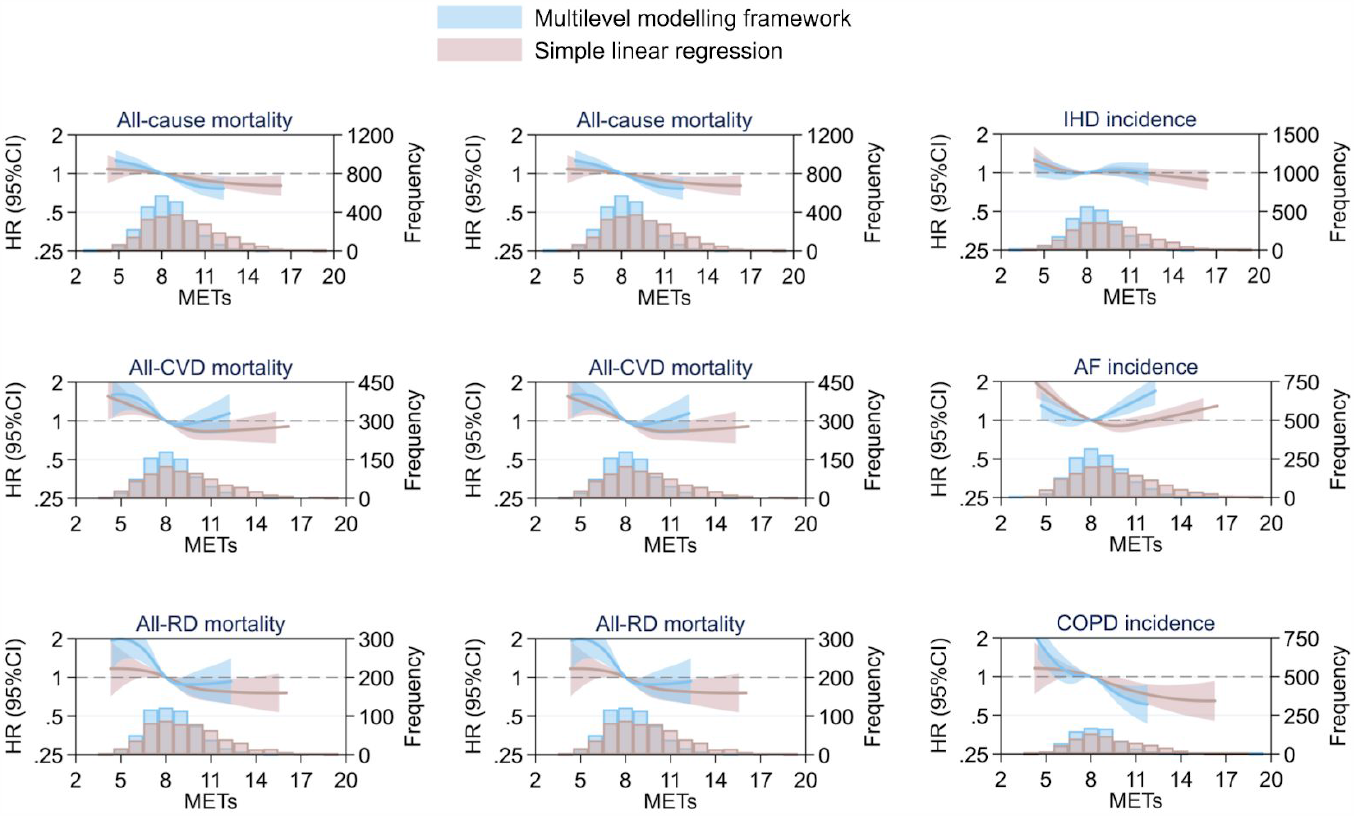
Sample-matched hazard ratios (HR) and 95% confidence intervals (CI) for nonlinear associations between fatal and non-fatal outcomes in the UK Biobank with cardiorespiratory fitness in metabolic equivalents (METs, per 3.5 ml O2·kg^-1^·min^-1^) estimated from the multilevel modelling framework and simple linear regression. Hazard ratios were computed relative to a fitness reference point of 8.0 METs. AF: atrial fibrillation; COPD: chronic obstructive pulmonary disease; CVD: cardiovascular disease; IHD: ischaemic heart disease; RD: respiratory disease. For these analyses, the analytical sample was matched between fitness estimation methods (exposure distributions shown by event status in superimposed histograms).

**Supplementary Figure 10.**
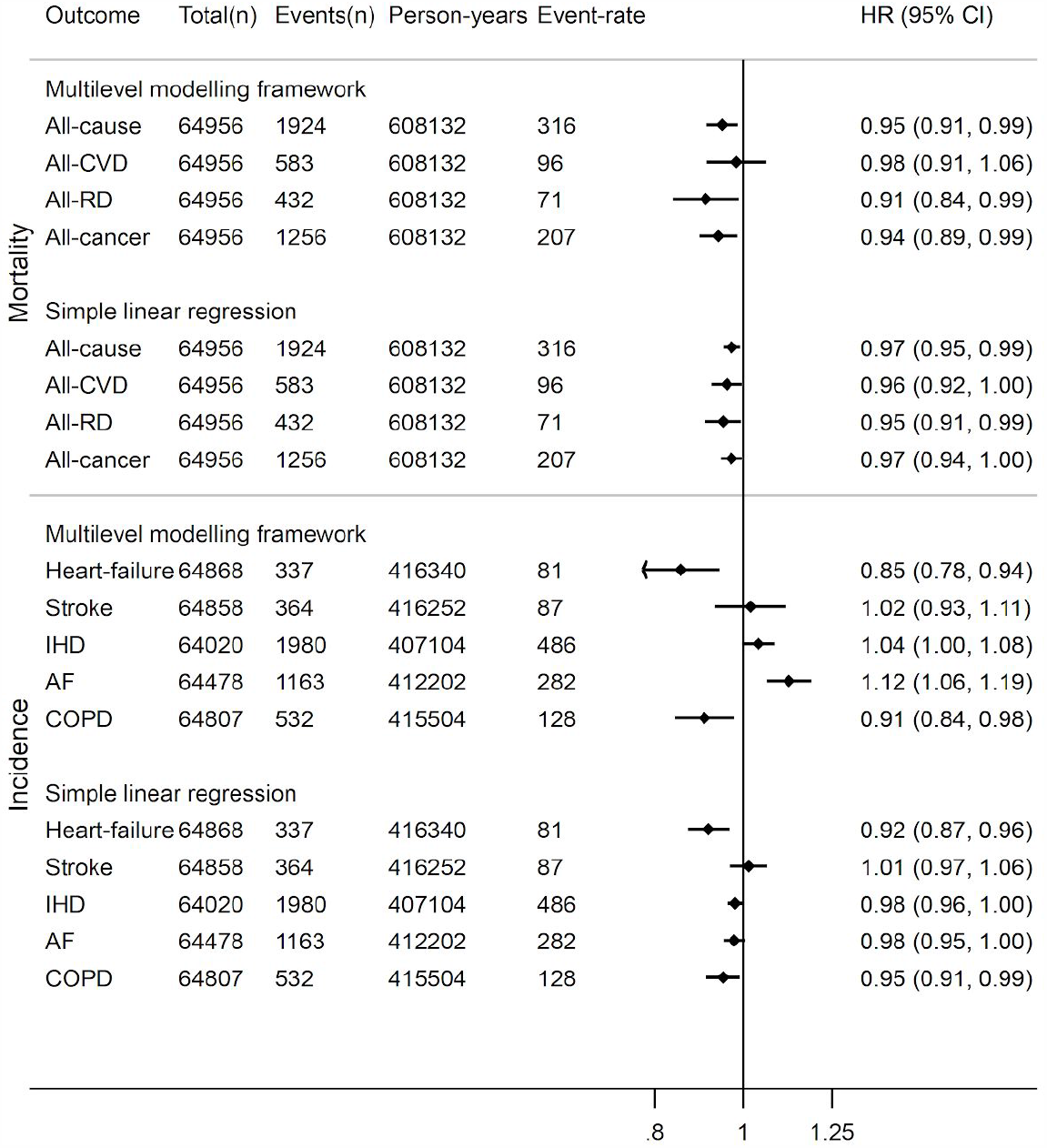
Sample-matched hazard ratios (HR) and 95% confidence intervals (CI) for prospective log-linear associations between fatal and non-fatal outcomes in the UK Biobank with cardiorespiratory fitness in metabolic equivalents (METs, per 3.5 ml O_2_·kg^-1^·min^-1^) estimated *using only model M5* from the multilevel modelling framework and the simple linear regression method. For these analyses, the analytical sample was matched between fitness estimation methods. Event rate per 100,000 person-years. AF - atrial fibrillation; COPD: chronic obstructive pulmonary disease; CVD: cardiovascular disease; IHD: ischaemic heart disease; RD-respiratory disease. COPD incidence mostly represents severe COPD since only ∼25% of cases end up in hospital.

**Supplementary Figure 11.**
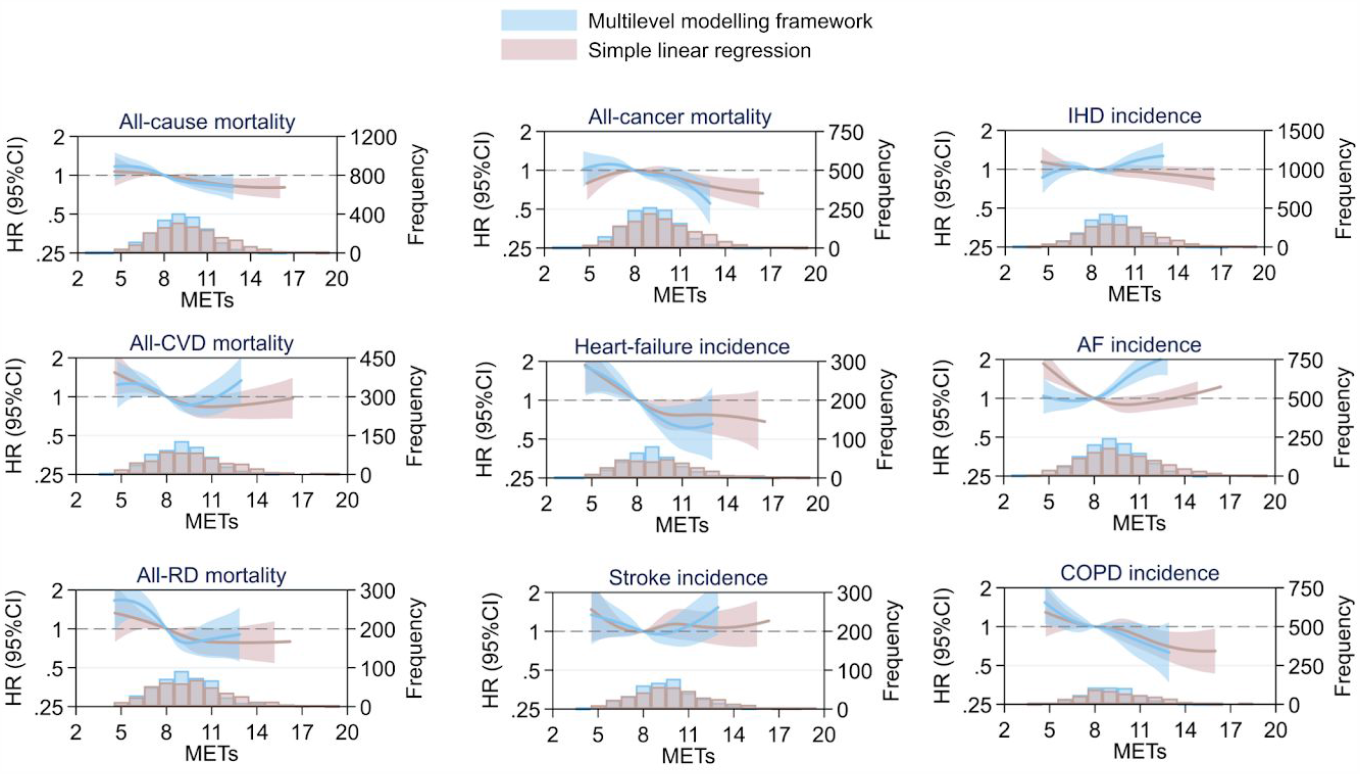
Hazard ratios (HR) and 95% confidence intervals (CI) for nonlinear (cubic spline) associations between fatal and non-fatal outcomes in the UK Biobank with cardiorespiratory fitness in metabolic equivalents (METs, per 3.5 ml O2·kg^-1^·min^-1^) estimated *using only model M5* from the multilevel modelling framework and the simple linear regression method. For these analyses, the analytical sample was matched between fitness estimation methods. (exposure distributions shown by event status in superimposed histograms). Hazard ratios were computed relative to a fitness reference point of 8.0 METs. AF: atrial fibrillation; COPD: chronic obstructive pulmonary disease; CVD: cardiovascular disease; IHD: ischaemic heart disease; RD: respiratory disease.

## Supplemental Tables

**Supplementary Table 1.** Sampling strata for validation study participants. Participants were selected using a stratified random sampling procedure for which the strata were sex, age (40-49y, 50-59y, 60-69y), and BMI (Supplemental Table 1). The range of each BMI strata covered at least the 25^th^ and 75^th^ percentile in the UKB sample, aiming to ensure that the validation study sample was broadly representative of fitness levels across strata in the UKB cohort.

| Age range (y)<br>Sex | 40-49 |  | 50-59 |  | 60-69 |  |
| --- | --- | --- | --- | --- | --- | --- |
|  | F | M | F | M | F | M |
| BMI group 1 | 20.5-23.9 | 22.0-25.4 | 21.0-23.9 | 22.5-25.4 | 21.5-24.4 | 22.9-25.8 |
| BMI group 2 | 24.0-27.4 | 25.5-28.4 | 24.0-27.4 | 25.5-28.9 | 24.5-28.4 | 25.9-28.9 |
| BMI group 3 | 27.5-35.0 | 28.5-33.5 | 27.5-35.0 | 29.0-34.0 | 28.5-34.5 | 29.0-33.5 |

F: Female, M: Male, BMI: Body mass index

**Supplementary Table 2.** Overview of tests completed by validation study participants; tests were parameterised according to the participant’s individualised UKB protocol. For example, a male participant with UKB test “M100” completed a flat test at 40W, two ramped tests with target WR values of 100W and 130W, a steady-state test, and a ramped VO2max test. Flat tests consisted of one steady-state work rate for 6 minutes. Ramped tests consisted of an initial steady-state WR for 2 minutes and incremented at a rate equal to RR for 4 minutes until the target WR was reached. Steady-state tests consisted of four consecutive steady-state work rates (WR1-4) at 4 minutes each. Maximal ramped tests consisted of an initial WR and incremented at a rate equal to RR until exhaustion.

| UKB allocation | Flat test | Low ramped test |  |  | High ramped test |  |  | Steady-state test |  |  |  | Ramped VO<sub>2</sub>max test |  |
| --- | --- | --- | --- | --- | --- | --- | --- | --- | --- | --- | --- | --- | --- |
|  | WR | Initial WR | Target WR | RR | Initial WR | Target WR | RR | WR 1 | WR 2 | WR 3 | WR 4 | Initial WR | RR |
| F30 | 30 | 30 | 50 | 5 | 30 | 80 | 12.5 | 45 | 55 | 65 | 75 | 65 | 15 |
| F40 | 30 | 30 | 50 | 5 | 30 | 80 | 12.5 | 45 | 55 | 65 | 75 | 65 | 15 |
| F50 | 30 | 30 | 50 | 5 | 30 | 80 | 12.5 | 45 | 55 | 65 | 75 | 65 | 15 |
| F60 | 30 | 30 | 60 | 7.5 | 30 | 90 | 15 | 45 | 55 | 65 | 75 | 65 | 15 |
| F70 | 30 | 30 | 70 | 10 | 30 | 100 | 17.5 | 45 | 55 | 65 | 75 | 65 | 15 |
| F80 | 30 | 30 | 80 | 12.5 | 30 | 100 | 17.5 | 45 | 55 | 65 | 75 | 65 | 15 |
| F90 | 30 | 30 | 60 | 7.5 | 30 | 90 | 15 | 45 | 60 | 75 | 90 | 75 | 20 |
| F100 | 30 | 30 | 70 | 10 | 30 | 100 | 17.5 | 45 | 60 | 75 | 90 | 75 | 20 |
| F110 | 30 | 30 | 70 | 10 | 30 | 110 | 17.5 | 45 | 60 | 75 | 90 | 75 | 20 |
| F120 | 30 | 30 | 70 | 10 | 30 | 110 | 20 | 45 | 60 | 75 | 90 | 75 | 20 |
| F130 | 30 | 30 | 70 | 10 | 30 | 110 | 20 | 45 | 60 | 75 | 90 | 75 | 20 |
| M40 | 40 | 40 | 70 | 7.5 | 40 | 110 | 17.5 | 60 | 75 | 90 | 105 | 90 | 20 |
| M50 | 40 | 40 | 70 | 7.5 | 40 | 110 | 17.5 | 60 | 75 | 90 | 105 | 90 | 20 |
| M60 | 40 | 40 | 70 | 7.5 | 40 | 110 | 17.5 | 60 | 75 | 90 | 105 | 90 | 20 |
| M70 | 40 | 40 | 70 | 7.5 | 40 | 110 | 17.5 | 60 | 75 | 90 | 105 | 90 | 20 |
| M80 | 40 | 40 | 80 | 10 | 40 | 120 | 20 | 60 | 75 | 90 | 105 | 90 | 20 |
| M90 | 40 | 40 | 90 | 12.5 | 40 | 130 | 22.5 | 60 | 75 | 90 | 105 | 90 | 20 |
| M100 | 40 | 40 | 100 | 15 | 40 | 130 | 22.5 | 60 | 75 | 90 | 105 | 90 | 20 |
| M110 | 40 | 40 | 80 | 10 | 40 | 110 | 17.5 | 60 | 80 | 100 | 120 | 100 | 30 |
| M120 | 40 | 40 | 90 | 12.5 | 40 | 120 | 20 | 60 | 80 | 100 | 120 | 100 | 30 |
| M130 | 40 | 40 | 100 | 15 | 40 | 130 | 22.5 | 60 | 80 | 100 | 120 | 100 | 30 |
| M140 | 40 | 40 | 100 | 15 | 40 | 140 | 25 | 60 | 80 | 100 | 120 | 100 | 30 |

UKB: UK Biobank, F: Female, M: Male, WR: Work rate (W), RR: Ramp rate (W·min<sup>-1</sup>)

**Supplementary Table 3.** Descriptions and coefficient estimates for the work rate estimation equations derived from multilevel modeling framework. Descriptions indicate the UKB CRF test phases used to compute features that are included as predictors for each equation.

| Equation Level | Source of information | Work rate estimation equation |
| --- | --- | --- |
| M1 | Ramp & recovery phases | $-55.5 + 1.42 \cdot HR_{rec45} + 0.567 \cdot b_0 + 14.8 \cdot RR^{0.5} - 1.14 \cdot HR_{rest} - 8.21 \cdot sex +$ $HR_{max} \cdot (1.11 - 0.0129 \cdot HR_{rec45} + 0.440 \cdot b_1 - 0.126 \cdot RR^{0.5} + 0.00693 \cdot HR_{rest} + 0.294 \cdot sex)$ |
| M2 | Recovery phase | $-66.8 + 0.523 \cdot HR_{rec45} + 2.54 \cdot HR_{rec0} - 274 \cdot RR^{0.5} - 3.58 \cdot HR_{rest} - 7.09 \cdot sex +$ $HR_{max} \cdot (2.16 - 0.00539 \cdot HR_{rec45} - 0.0252 \cdot HR_{rec0} + 2.74 \cdot RR^{0.5} + 0.0197 \cdot HR_{rest} + 0.360 \cdot sex)$ |
| M3 | Ramp phase | $-61.2 + 0.626 \cdot b_0 + 89.0 \cdot RR^{0.5} + 0.394 \cdot HR_{rest} - 12.3 \cdot sex +$ $HR_{max} \cdot (1.07 + 0.487 \cdot b_1 - 0.806 \cdot RR^{0.5} - 0.00589 \cdot HR_{rest} + 0.319 \cdot sex)$ |
| M4 | Flat & recovery phases | $-13.9 - 1.05 \cdot HR_{rec45} + 0.153 \cdot HR_{flat} - 45.0 \cdot sex +$ $HR_{max} \cdot (2.32 + 0.0101 \cdot HR_{rec45} - 0.0209 \cdot HR_{flat} + 0.687 \cdot sex)$ |
| M5 | Flat phase (first 2 min) | $-10.5 - 1.03 \cdot HR_{rest} - 0.0233 \cdot HR_{flat} - 47.1 \cdot sex +$ $HR_{max} \cdot (2.30 + 0.0121 \cdot HR_{rest} - 0.0210 \cdot HR_{flat} + 0.707 \cdot sex)$ |

$HR_{max}$ : Maximal heart rate (either age-predicted or directly measured), $HR_{rest}$ : Resting heart rate, $HR_{rec45}$ : Recovery heart rate at 45s post-exercise, $HR_{rec0}$ : Recovery heart rate at 0s post-exercise, $b_0$ : Intercept from the ramp phase linear regression model, $b_1$ : Slope from the ramp phase linear regression model, $HR_{flat}$ : Median heart rate computed for the flat phase, $RR^{0.5}$ : Square root of test ramp rate, $sex$ : “0” females, “1” males

**Supplementary Table 4.**
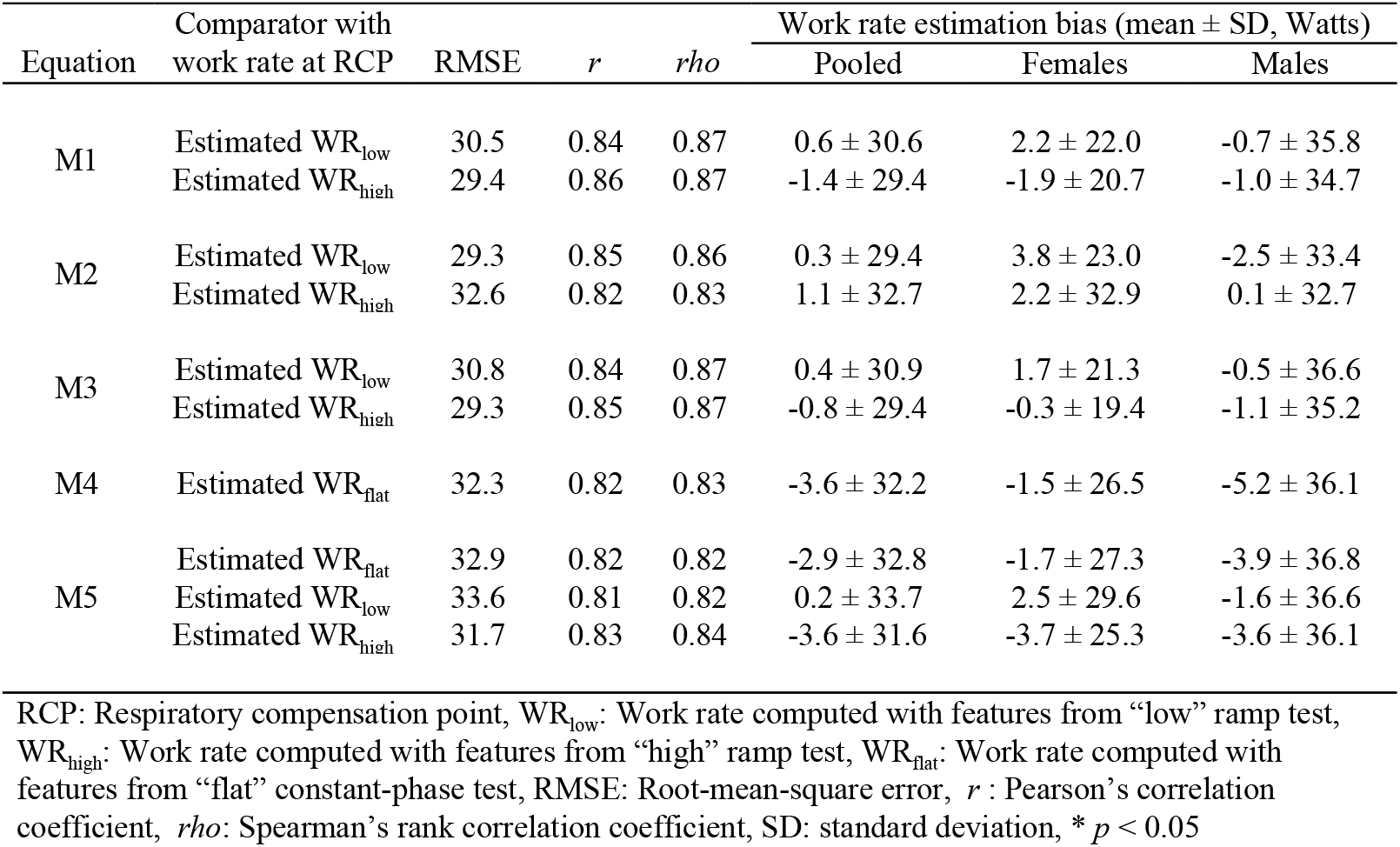
Agreement between work rates measured at the respiratory compensation point (RCP, see Supplemental Figure 1) and work rates estimated from the flat, low, and high ramp exercise tests in the validation study. M1 results are shown in Figure 2.

| Equation | Comparator with work rate at RCP | RMSE | *r* | *rho* | Work rate estimation bias (mean ± SD, Watts) | | |
| --- | --- | --- | --- | --- | --- | --- | --- |
|  |  |  |  |  | Pooled | Females | Males |
| M1 | Estimated WR<sub>low</sub> | 30.5 | 0.84 | 0.87 | 0.6 ± 30.6 | 2.2 ± 22.0 | -0.7 ± 35.8 |
| | Estimated WR<sub>high</sub> | 29.4 | 0.86 | 0.87 | -1.4 ± 29.4 | -1.9 ± 20.7 | -1.0 ± 34.7 |
| M2 | Estimated WR<sub>low</sub> | 29.3 | 0.85 | 0.86 | 0.3 ± 29.4 | 3.8 ± 23.0 | -2.5 ± 33.4 |
| | Estimated WR<sub>high</sub> | 32.6 | 0.82 | 0.83 | 1.1 ± 32.7 | 2.2 ± 32.9 | 0.1 ± 32.7 |
| M3 | Estimated WR<sub>low</sub> | 30.8 | 0.84 | 0.87 | 0.4 ± 30.9 | 1.7 ± 21.3 | -0.5 ± 36.6 |
| | Estimated WR<sub>high</sub> | 29.3 | 0.85 | 0.87 | -0.8 ± 29.4 | -0.3 ± 19.4 | -1.1 ± 35.2 |
| M4 | Estimated WR<sub>flat</sub> | 32.3 | 0.82 | 0.83 | -3.6 ± 32.2 | -1.5 ± 26.5 | -5.2 ± 36.1 |
| M5 | Estimated WR<sub>flat</sub> | 32.9 | 0.82 | 0.82 | -2.9 ± 32.8 | -1.7 ± 27.3 | -3.9 ± 36.8 |
| | Estimated WR<sub>low</sub> | 33.6 | 0.81 | 0.82 | 0.2 ± 33.7 | 2.5 ± 29.6 | -1.6 ± 36.6 |
| | Estimated WR<sub>high</sub> | 31.7 | 0.83 | 0.84 | -3.6 ± 31.6 | -3.7 ± 25.3 | -3.6 ± 36.1 |

RCP: Respiratory compensation point, WR<sub>low</sub>: Work rate computed with features from “low” ramp test, WR<sub>high</sub>: Work rate computed with features from “high” ramp test, WR<sub>flat</sub>: Work rate computed with features from “flat” constant-phase test, RMSE: Root-mean-square error, *r*: Pearson’s correlation coefficient, *rho*: Spearman’s rank correlation coefficient, SD: standard deviation, \* $p < 0.05$

**Supplementary Table 5.** Agreement between work rates measured at the lactate threshold (LT, see Supplemental Figure 1) and work rates estimated from the flat, low, and high ramp exercise tests in the validation study.

| Equation | Comparator with<br>work rate at LT | RMSE | *r* | *rho* | Work rate estimation bias (mean ± SD, Watts) | | |
| --- | --- | --- | --- | --- | --- | --- | --- |
|  |  |  |  |  | Pooled | Females | Males |
| M1 | Estimated WR<sub>low</sub> | 58.0 | 0.83 | 0.86 | 49.8 ± 29.9* | 37.1 ± 19.7* | 60.5 ± 32.8* |
| | Estimated WR<sub>high</sub> | 55.9 | 0.86 | 0.88 | 47.4 ± 29.8* | 32.6 ± 20.0* | 60.0 ± 31.0* |
| M2 | Estimated WR<sub>low</sub> | 59.4 | 0.80 | 0.82 | 50.4 ± 31.4* | 40.7 ± 32.5* | 59.2 ± 27.9* |
| | Estimated WR<sub>high</sub> | 62.2 | 0.77 | 0.79 | 50.7 ± 36.1* | 39.2 ± 41.0* | 61.3 ± 27.1* |
| M3 | Estimated WR<sub>low</sub> | 57.6 | 0.84 | 0.86 | 49.7 ± 29.1* | 36.9 ± 18.1* | 60.6 ± 32.2* |
| | Estimated WR<sub>high</sub> | 55.5 | 0.86 | 0.88 | 48.3 ± 27.5* | 34.7 ± 16.2* | 59.9 ± 29.9* |
| M4 | Estimated WR<sub>flat</sub> | 56.2 | 0.83 | 0.84 | 47.8 ± 29.5* | 36.0 ± 24.2* | 58.5 ± 30.0* |
| M5 | Estimated WR<sub>flat</sub> | 56.1 | 0.83 | 0.83 | 47.9 ± 29.2* | 36.2 ± 23.9* | 58.5 ± 29.7* |
| | Estimated WR<sub>low</sub> | 58.9 | 0.82 | 0.83 | 51.0 ± 29.5* | 39.3 ± 26.1* | 61.3 ± 28.6* |
| | Estimated WR<sub>high</sub> | 55.5 | 0.84 | 0.85 | 47.5 ± 28.8* | 34.1 ± 22.8* | 59.6 ± 28.4* |

LT: lactate threshold, WR<sub>low</sub>: Work rate computed with features from “low” ramp test, WR<sub>high</sub>: Work rate computed with features from “high” ramp test, WR<sub>flat</sub>: Work rate computed with features from “flat” constant-phase test, RMSE: Root-mean-square error, *r*: Pearson’s correlation coefficient, *rho*: Spearman’s rank correlation coefficient, SD: standard deviation, \* $p < 0.05$

**Supplementary Table 6.**
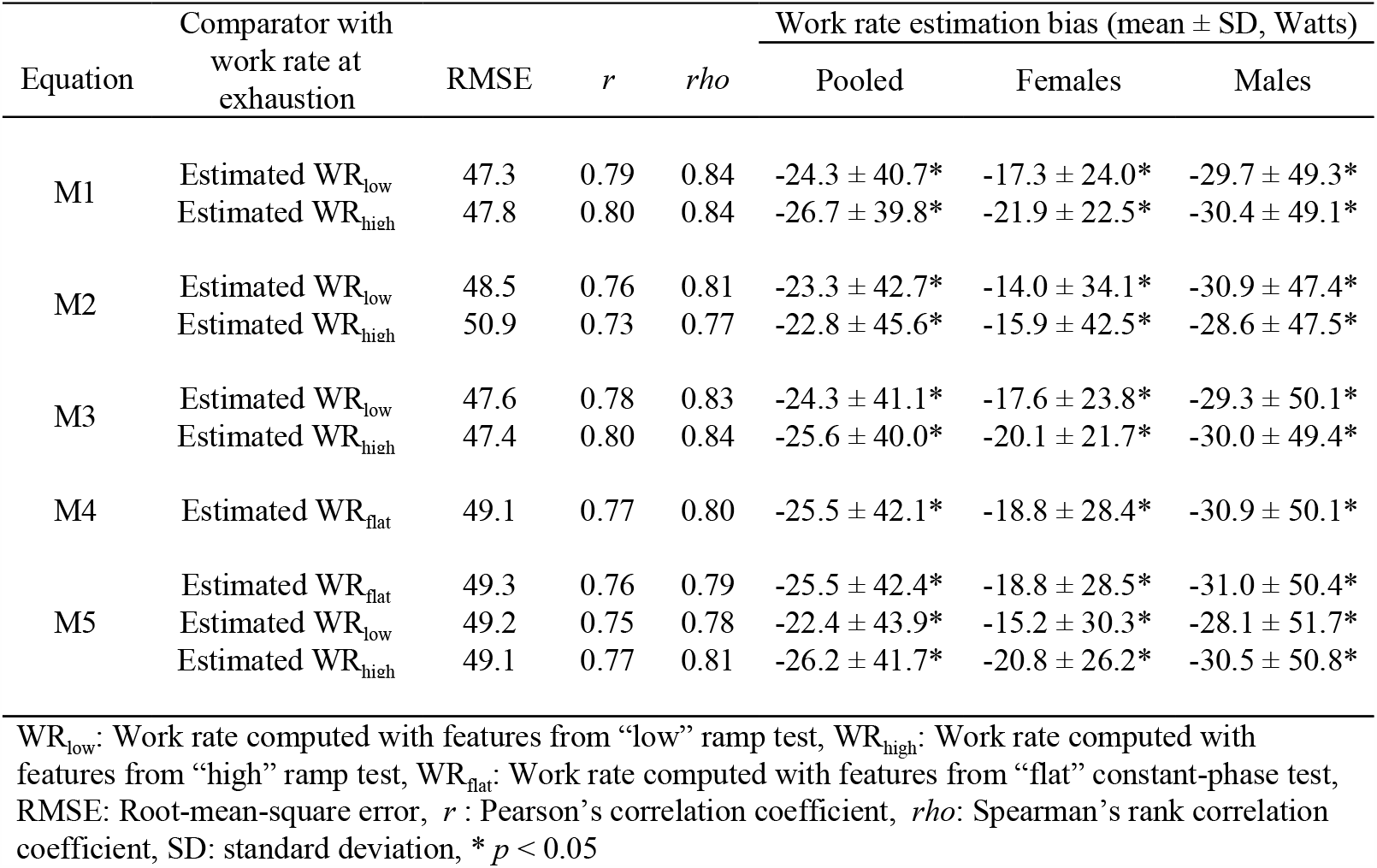
Agreement between work rates measured at exhaustion and work rates computed from work rates estimated from the flat, low, and high ramp exercise tests in the validation study.

**Supplementary Table 7.** Agreement between directly measured at VO_2_max and VO_2_max values computed from different exercise tests and work rate estimation equations, using age-predicted maximal heart rate. M1 results are shown in Figure 3.

| Equation | Comparator with measured $\text{VO}_2\text{max}$ | RMSE | $r$ | $\rho$ | VO<sub>2</sub>max estimation bias (mean ± SD, ml O<sub>2</sub>·kg<sup>-1</sup>·min<sup>-1</sup>) | | |
| --- | --- | --- | --- | --- | --- | --- | --- |
|  |  |  |  |  | Pooled | Females | Males |
| M1 | Estimated $\text{VO}_2\text{max}_{\text{low}}$ | 4.9 | 0.70 | 0.74 | 0.1 ± 4.9 | -0.1 ± 4.4 | 0.2 ± 5.3 |
| | Estimated $\text{VO}_2\text{max}_{\text{high}}$ | 4.8 | 0.72 | 0.74 | -0.2 ± 4.8 | -0.6 ± 4.4 | 0.0 ± 5.1 |
| M2 | Estimated $\text{VO}_2\text{max}_{\text{low}}$ | 4.5 | 0.74 | 0.74 | -0.1 ± 4.6 | -0.1 ± 4.6 | -0.1 ± 4.5 |
| | Estimated $\text{VO}_2\text{max}_{\text{high}}$ | 4.7 | 0.73 | 0.72 | -0.2 ± 4.7 | -0.8 ± 4.8 | 0.2 ± 4.6 |
| M3 | Estimated $\text{VO}_2\text{max}_{\text{low}}$ | 5.0 | 0.68 | 0.74 | 0.0 ± 5.0 | -0.3 ± 4.4 | 0.3 ± 5.5 |
| | Estimated $\text{VO}_2\text{max}_{\text{high}}$ | 4.8 | 0.70 | 0.73 | -0.1 ± 4.8 | -0.5 ± 4.4 | 0.1 ± 5.2 |
| M4 | Estimated $\text{VO}_2\text{max}_{\text{flat}}$ | 5.0 | 0.68 | 0.68 | -0.3 ± 5.0 | -0.4 ± 5.1 | -0.2 ± 4.9 |
| M5 | Estimated $\text{VO}_2\text{max}_{\text{low}}$ | 4.9 | 0.69 | 0.68 | -0.3 ± 4.9 | -0.3 ± 5.0 | -0.3 ± 4.9 |
| | Estimated $\text{VO}_2\text{max}_{\text{high}}$ | 4.8 | 0.70 | 0.70 | 0.2 ± 4.9 | 0.4 ± 4.9 | -0.1 ± 4.9 |
| | Estimated $\text{VO}_2\text{max}_{\text{flat}}$ | 4.8 | 0.71 | 0.70 | -0.3 ± 4.8 | -0.2 ± 4.8 | -0.3 ± 4.8 |

VO<sub>2</sub>max: Maximal oxygen consumption, $\text{VO}_2\text{max}_{\text{low}}$ : $\text{VO}_2\text{max}$ computed with features from “low” ramp test, $\text{VO}_2\text{max}_{\text{high}}$ : $\text{VO}_2\text{max}$ computed with features from “high” ramp test, $\text{VO}_2\text{max}_{\text{flat}}$ : $\text{VO}_2\text{max}$ computed with features from “flat” constant-phase test, RMSE: Root-mean-square error, $r$ : Pearson’s correlation coefficient, $\rho$ : Spearman’s rank correlation coefficient, SD: standard deviation, \* $p < 0.05$

**Supplementary Table 8.** Agreement between VO_2_max estimated within each level of the set of work rate estimation equations for M1-M3 and M5 when using features computed from ramp tests, and between M4 and M5 when using features computed from flat tests. Bias values were computed as the difference between the first and second comparators.

| First comparator | Second comparator | RMSE | $r$ | $\rho$ | VO<sub>2</sub>max estimation bias (mean ± SD,<br>ml O<sub>2</sub>·kg<sup>-1</sup>·min<sup>-1</sup>) | | |
| --- | --- | --- | --- | --- | --- | --- | --- |
|  |  |  |  |  | Pooled | Females | Males |
| VO<sub>2</sub>max<sub>high</sub> from M1 | VO<sub>2</sub>max<sub>low</sub> from M1 | 2.2 | 0.94 | 0.94 | -0.4 ± 2.2* | -0.5 ± 2.3 | -0.2 ± 1.9 |
| VO<sub>2</sub>max<sub>high</sub> from M2 | VO<sub>2</sub>max<sub>low</sub> from M2 | 3.1 | 0.96 | 0.96 | 0.0 ± 3.1 | -0.7 ± 2.0* | 0.2 ± 1.1 |
| VO<sub>2</sub>max<sub>high</sub> from M3 | VO<sub>2</sub>max<sub>low</sub> from M3 | 1.9 | 0.94 | 0.95 | -0.2 ± 1.9 | -0.2 ± 1.8 | -0.2 ± 1.9 |
| VO<sub>2</sub>max<sub>flat</sub> from M4 | VO<sub>2</sub>max<sub>flat</sub> from M5 | 0.9 | 0.99 | 0.99 | -0.0 ± 0.9 | -0.1 ± 0.8 | 0.0 ± 0.9 |
| VO<sub>2</sub>max<sub>high</sub> from M5 | VO<sub>2</sub>max<sub>low</sub> from M5 | 1.9 | 0.98 | 0.98 | -0.6 ± 1.8* | -0.6 ± 1.0* | -0.3 ± 0.9* |

VO<sub>2</sub>max: Maximal oxygen consumption, VO<sub>2</sub>max<sub>low</sub>: VO<sub>2</sub>max computed with features from “low” ramp test, VO<sub>2</sub>max<sub>high</sub>: VO<sub>2</sub>max computed with features from “high” ramp test, VO<sub>2</sub>max<sub>flat</sub>: VO<sub>2</sub>max computed with features from “flat” constant-phase test, RMSE: Root-mean-square error, $r$ : Pearson’s correlation coefficient, $\rho$ : Spearman’s rank correlation coefficient, SD: standard deviation, \* $p < 0.05$

**Supplementary Table 9.** Agreement between directly measured at VO_2_max and VO_2_max values estimated from different exercise tests and work rate estimated equations, using directly measured maximal heart rate.

| Equation | Comparator with measured $\text{VO}_2\text{max}$ | RMSE | $r$ | $\rho$ | $\text{VO}_2\text{max}$ estimation bias (mean ± SD, ml $\text{O}_2 \cdot \text{kg}^{-1} \cdot \text{min}^{-1}$) | | |
| --- | --- | --- | --- | --- | --- | --- | --- |
|  |  |  |  |  | Pooled | Females | Males |
| M1 | Estimated $\text{VO}_2\text{max}_{\text{low}}$ | 4.8 | 0.71 | 0.74 | $0.3 \pm 4.8$ | $0.3 \pm 4.2$ | $0.3 \pm 5.2$ |
| | Estimated $\text{VO}_2\text{max}_{\text{high}}$ | 4.4 | 0.75 | 0.75 | $-0.1 \pm 4.5$ | $-0.2 \pm 4.3$ | $0.0 \pm 4.6$ |
| M2 | Estimated $\text{VO}_2\text{max}_{\text{low}}$ | 4.8 | 0.70 | 0.69 | $0.2 \pm 4.8$ | $0.3 \pm 4.5$ | $0.1 \pm 5.1$ |
| | Estimated $\text{VO}_2\text{max}_{\text{high}}$ | 4.7 | 0.72 | 0.70 | $-0.1 \pm 4.7$ | $-0.5 \pm 4.5$ | $0.3 \pm 4.8$ |
| M3 | Estimated $\text{VO}_2\text{max}_{\text{low}}$ | 4.9 | 0.69 | 0.71 | $0.3 \pm 4.9$ | $0.1 \pm 4.2$ | $0.4 \pm 5.4$ |
| | Estimated $\text{VO}_2\text{max}_{\text{high}}$ | 4.5 | 0.74 | 0.74 | $0.1 \pm 4.6$ | $-0.1 \pm 4.3$ | $0.2 \pm 4.8$ |
| M4 | Estimated $\text{VO}_2\text{max}_{\text{flat}}$ | 4.9 | 0.69 | 0.67 | $0.0 \pm 4.9$ | $0.0 \pm 4.8$ | $0.0 \pm 5.1$ |
| M5 | Estimated $\text{VO}_2\text{max}_{\text{low}}$ | 4.9 | 0.69 | 0.67 | $0.0 \pm 4.9$ | $0.1 \pm 4.7$ | $0.0 \pm 5.1$ |
| | Estimated $\text{VO}_2\text{max}_{\text{high}}$ | 5.0 | 0.68 | 0.68 | $0.5 \pm 5.0$ | $0.9 \pm 4.7$ | $0.1 \pm 5.3$ |
| | Estimated $\text{VO}_2\text{max}_{\text{flat}}$ | 5.0 | 0.68 | 0.67 | $0.0 \pm 5.0$ | $0.2 \pm 4.7$ | $-0.1 \pm 5.3$ |

$\text{VO}_2\text{max}$ : Maximal oxygen consumption, $\text{VO}_2\text{max}_{\text{low}}$ : $\text{VO}_2\text{max}$ computed with features from “low” ramp test, $\text{VO}_2\text{max}_{\text{high}}$ : $\text{VO}_2\text{max}$ computed with features from “high” ramp test, $\text{VO}_2\text{max}_{\text{flat}}$ : $\text{VO}_2\text{max}$ computed with features from “flat” constant-phase test, RMSE: Root-mean-square error, $r$ : Pearson’s correlation coefficient, $\rho$ : Spearman’s rank correlation coefficient, SD: standard deviation, \* $p < 0.05$

## Notes

### Competing Interest Statement

The authors have declared no competing interest.

### Author Declarations

University of Cambridge Human Biology Research Ethics Committee (Ref: HBREC/2015.16)

